# Persistent immune dysregulation and metabolic alterations following SARS-CoV-2 infection

**DOI:** 10.1101/2025.04.16.25325949

**Authors:** Silvia Lucena Lage, Katherine Bricker-Holt, Joseph M. Rocco, Adam Rupert, Frank X. Donovan, Yevgeniya A. Abramzon, Settara C. Chandrasekharappa, Colton McNinch, Logan Cook, Eduardo Pinheiro Amaral, Gabriel Rosenfeld, Thomas Dalhuisen, Avery Eun, Rebecca Hoh, Emily Fehrman, Jeffrey N. Martin, Steven G. Deeks, Timothy J. Henrich, Michael J. Peluso, Irini Sereti

## Abstract

SARS-CoV-2 can cause a variety of post-acute sequelae including Long COVID19 (LC), a complex, multisystem disease characterized by a broad range of symptoms including fatigue, cognitive impairment, and post-exertional malaise. The pathogenesis of LC is incompletely understood. In this study, we performed comprehensive cellular and transcriptional immunometabolic profiling within a cohort that included SARS-CoV-2-naïve controls (NC, N=30) and individuals with prior COVID-19 (∼4-months) who fully recovered (RC, N=38) or went on to experience Long COVID symptoms (N=58). Compared to the naïve controls, those with prior COVID-19 demonstrated profound metabolic and immune alterations at the proteomic, cellular, and epigenetic level. Specifically, there was an enrichment in immature monocytes with sustained inflammasome activation and oxidative stress, elevated arachidonic acid levels, decreased tryptophan, and variation in the frequency and phenotype of peripheral T-cells. Those with LC had increased CD8 T-cell senescence and a distinct transcriptional profile within CD4 and CD8 T-cells and monocytes by single cell RNA sequencing. Our findings support a profound and persistent immunometabolic dysfunction that follows SARS-CoV-2 which may form the pathophysiologic substrate for LC. Our findings suggest that trials of therapeutics that help restore immune and metabolic homeostasis may be warranted to prevent, reduce, or resolve LC symptoms.

## INTRODUCTION

Long COVID (LC) is a multisystem condition following the acute phase of COVID-19, in which individuals experience persistent or new COVID-attributed symptoms including fatigue, cognitive impairment and post-exertional malaise (*1, 2*). LC has been documented to last for years following SARS-CoV-2 infection (*3, 4*), and a subset of individuals can be profoundly disabled. The proposed mechanisms of LC include immune dysregulation, viral persistence, microbial dysbiosis, autoimmunity, and dysfunctional neurological signaling (*1, 4, 5*).

Mechanisms such as oxidative stress and inflammation have been associated with chronic fatigue, depression, and anxiety following acute COVID-19 (*6, 7*). We previously showed that oxidative stress-associated inflammasome activation in circulating classical monocytes (CD14^high^CD16^-^) from COVID-19 patients persists after a short recovery period (*8*). Inflammasomes, formed in the cytosol in response to pathogen- or damage-associated molecular patterns (PAMPs/DAMPs), activate pro-caspase-1, which processes IL-1β and IL-18 into their active forms and coordinates their secretion (*9*). Oxidative stress mediators, such as reactive oxygen species (ROS) and toxic lipid peroxides, also trigger inflammasomes, creating a feedback loop that promotes a systemic hyperinflammatory state. This hyperinflammation leads to tissue damage and is being explored as a target for host-directed therapies to reduce/mitigate COVID-19 complications (*10*).

T-cells are also crucial in regulating COVID-19 pathogenesis, and disruptions in the adaptive immune system can affect acute responses to the virus. Delayed T-cell responses, caused by an uncoordinated innate and adaptive response, and sustained CD8 T-cell dysfunction have been linked to severe COVID-19 (*11*). Prolonged inflammation and T-cell activation, marked by exhaustion indicators like PD-1 and TIM-3, also predict worse outcomes during acute COVID-19 (*12*). Beyond the acute phase, improper adaptive immune responses contribute to LC, characterized by systemic inflammation, immune dysregulation, and abnormal CD4 and CD8 T-cell behavior (*13, 14*). T-cell exhaustion and immunosenescence have also been linked with LC (*15–17*). For example, a pilot study in individuals recovering from COVID-19 found a connection between CD57 expression, a marker of cellular senescence and extensive proliferative history (*18*), on CD8 T-cells and the persistence of symptoms three months following SARS-CoV-2 infection (*19*), but this has not been further evaluated.

Here we performed an extensive multi-omic analysis evaluating cytokines, lipid mediators, and reactive oxygen species as well as cellular immune, transcriptional, and epigenetic profiles in a cohort of participants approximately 4 months following their first episode of COVID-19 and investigated whether these responses associate with the presence of LC symptoms. We found pronounced alterations in many of these measurements months after the initial SARS-CoV-2 infection that were present even in those without persistent symptoms; we also identified perturbations in these biological pathways that were more evident among those with LC compared to those who fully recovered. Understanding the potential mechanisms underlying LC could guide management strategies aimed at addressing long-term symptoms or provide rationale for therapeutic trials aimed at preventing or reversing LC and other post-acute sequelae.

## RESULTS

### Subhead 1: Cohort characteristics

We analyzed blood samples collected at a median of 4.1 months following nucleic acid-confirmed SARS-CoV-2 infection in the pre-Omicron era from 96 individuals from the San Francisco-based Long-term Impact of Infection with Novel Coronavirus (LIINC) cohort study (NCT04362150). Cohort procedures have previously been described in detail (*20*). For comparison, we included 30 contemporaneous controls matched for age, sex, and race who had not experienced COVID-19 as confirmed also by negative serology and PCR testing, (no COVID-19, NC) from the Longitudinal Study of COVID-19 Sequalae and Immunity (RECON-19) cohort (NCT044411147). All study participants provided informed consent prior to any study procedures.

LIINC participants were further divided into individuals with Long COVID symptoms (LC, n=58) and those who reported full recovery (RC, n=38). Consistent with the case definition from the National Academies of Sciences Engingeering, and Medicine (NASEM) (*2*), LC was defined as the presence of COVID-attributed symptoms for 3 or more months following SARS-CoV-2 infection. Importantly, symptoms that pre-dated the onset of COVID-19 and did not worsen were not considered to represent LC. RC was defined as the absence of any COVID-attributed symptoms over the same period. The LC group had a median age of 47 years and was comprised of 63.8% females. The median age of the RC group was 47 years and 47.4% were female. 27.6% of the LC group had been hospitalized during the acute phase of COVID-19, compared to 18.4% of recovered patients. Full demographic and clinical data are available in Table 1 and Supplemental Table 1, respectively.

**Table 1.**
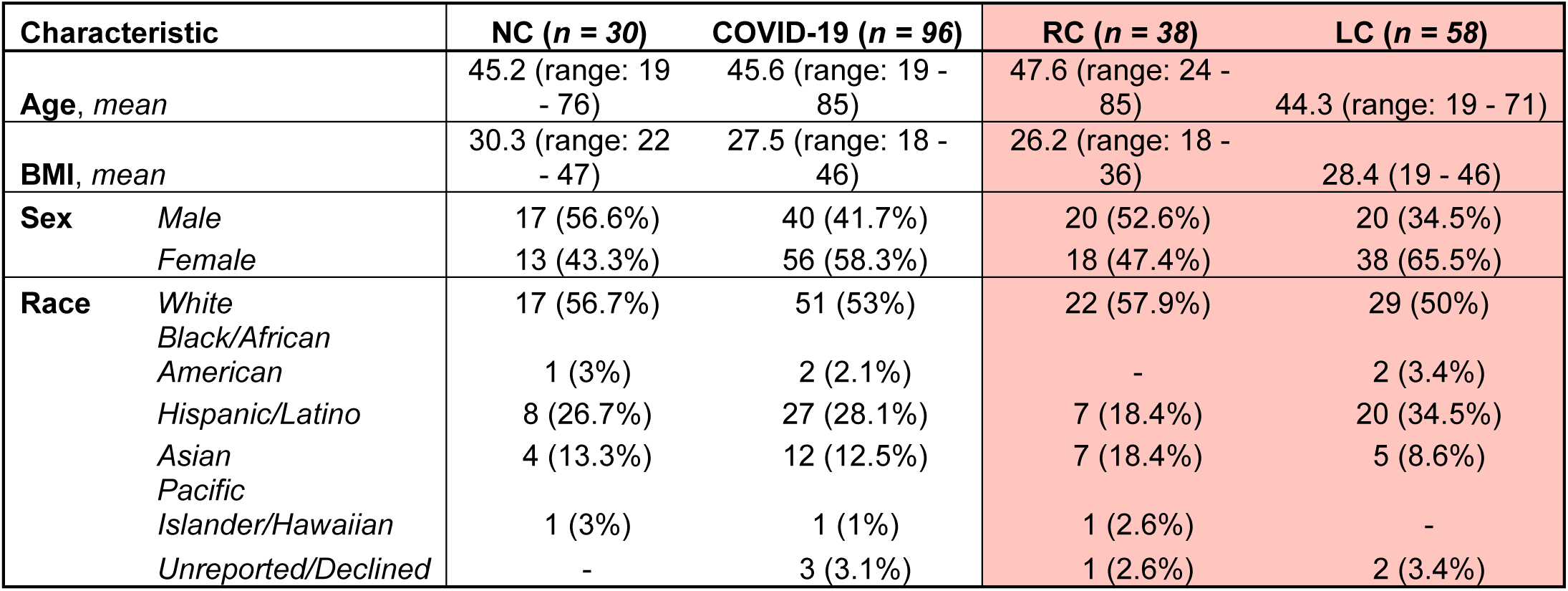
Demographic characteristics by SARS-CoV-2 infection status. NC, SARS-CoV-2 unexposed controls; RC, recovered COVID-19; LC, Long COVID symptoms.

### Subhead 2: Oxidative stress activity associates with immature monocytes in LC

Monocytes play a crucial role in COVID-19 immunopathogenesis, undergoing oxidative stress and releasing inflammatory mediators that contribute to hyperinflammation and disease severity (*8, 21–23*). In acute COVID-19, a reduction in non-classical monocytes (CD14^low^CD16^+^) with an enrichment of classical monocytes is observed (*8, 24*). In severe cases, dysfunctional HLA-DR^lo^CD163^hi^ monocytes also emerge (*8, 25*). While their role in acute COVID-19 is well-documented, research on monocytes in LC is very limited (*26, 27*).

Compared to SARS-CoV-2-naïve controls (NC), we found that individuals with prior SARS-CoV-2 infection had a decreased proportion of classical monocytes (Figure 1A) and similar percentages of intermediate (Figure 1B) and non-classical monocytes (Figure 1C). Despite equivalent CD163 expression levels (Figure 1D), monocytes from participants with prior COVID-19 had reduced expression of HLA-DR (Figure 1E) and p16^INK4a^, a marker of replicative senescence (Figure 1F), compared to NC, indicating the presence of immature circulating monocytes months after acute SARS-CoV-2 infection. To further evaluate whether this innate immune phenotype is associated with LC, we compared participants with LC symptoms (LC) with participants who fully recovered (RC) (Table 1). The proportion of monocyte subsets (Supplemental Figure 1A-C) and expression levels of CD163 (Supplemental Figure 1D), HLA-DR (Supplemental Figure 1E) and p16^INK4a^ (Supplemental Figure 1F) alone could not distinguish LC from RC participants.

**Figure 1.**
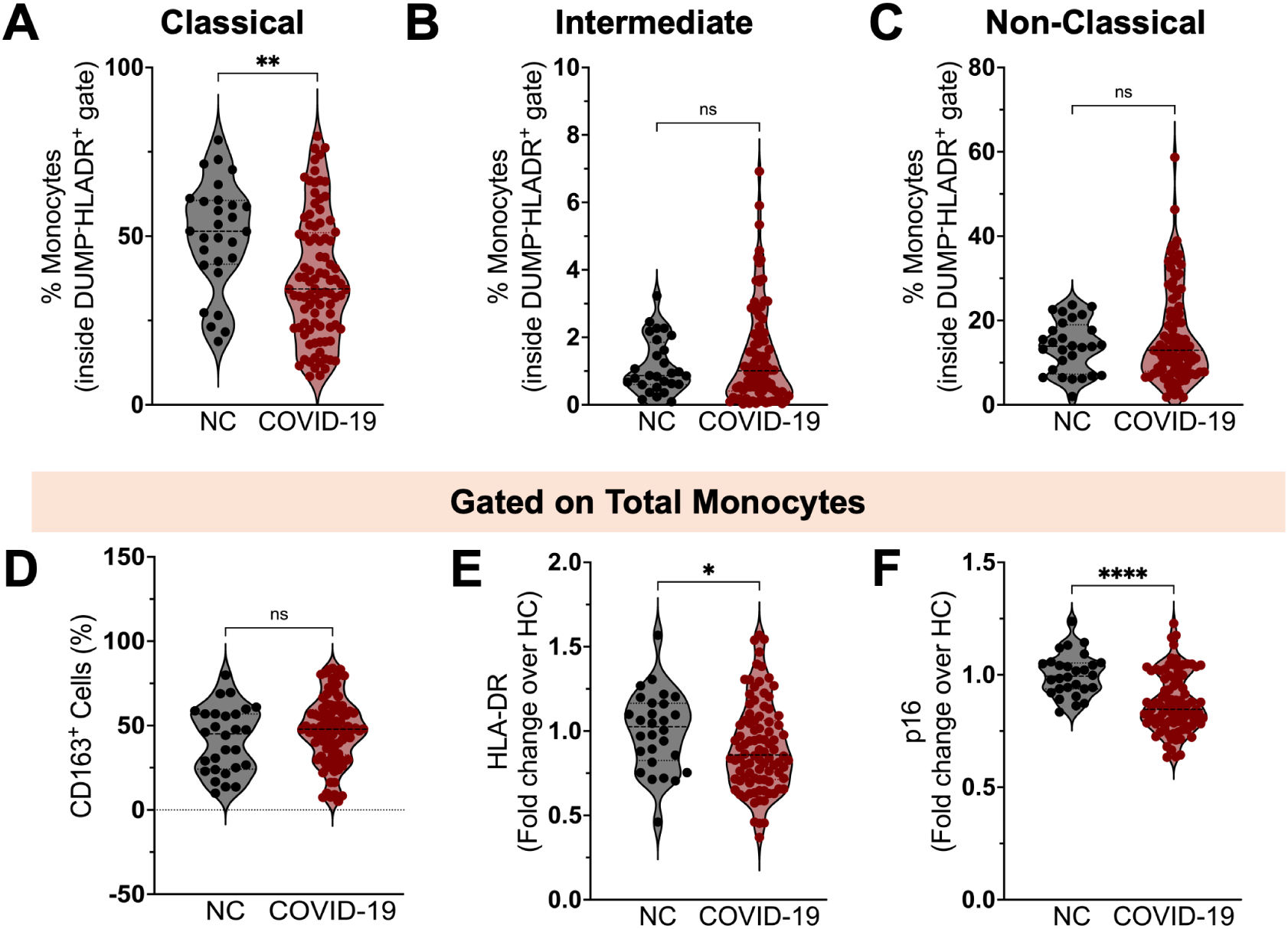
Phenotypic changes in circulating monocyte subsets post-COVID-19. Percentage of classical **(A)**, intermediate **(B)** and non-classical **(C)** monocytes among circulating mononuclear myeloid cells (HLADR+CD2−CD3−CD19−CD20−CD56−CD66−) in SARS-CoV-2-naïve controls (NC, n=28) and COVID-19 participants (n=92). Percentages of CD163^+^ cells **(D)**, and geometric mean fluorescence intensity (gMFI) expression of HLA-DR **(E)** and p16^INK4a^ **(F)** on circulating monocytes from NC and COVID-19 individuals. gMFIdata are expressed as median of the fold change (gMFI of each sample over the gMFI mean of the NC group). Bars represent median ± quartiles. Statistical analysis was performed using a two-sided Mann-Whitney test (*, *P* < 0.05, **, *P* < 0.005, ***, *P* < 0.0005).

Because of their importance in the pathophysiology of COVID-19, prolonged inflammasome activation and oxidative stress have been suggested as possible drivers of LC (*8, 10*). We assessed plasma levels of IL-1β and IL-18 as well as caspase-1/4/5 activity on circulating blood monocytes as readouts of systemic inflammasome activation. Compared to naïve controls, we observed higher caspase-1/4/5 activity (measured as gMFI of the caspase-1/4/5 fluorescent inhibitor FLICA) in monocytes from participants with prior COVID-19 (Figure 2A). This was accompanied by increased plasma levels of IL-1β (Figure 2B), suggesting systemic and persistent inflammasome activation in the post-acute phase. In contrast, IL-18 levels did not distinguish uninfected individuals from those with prior COVID-19 (Figure 2C).

**Figure 2.**
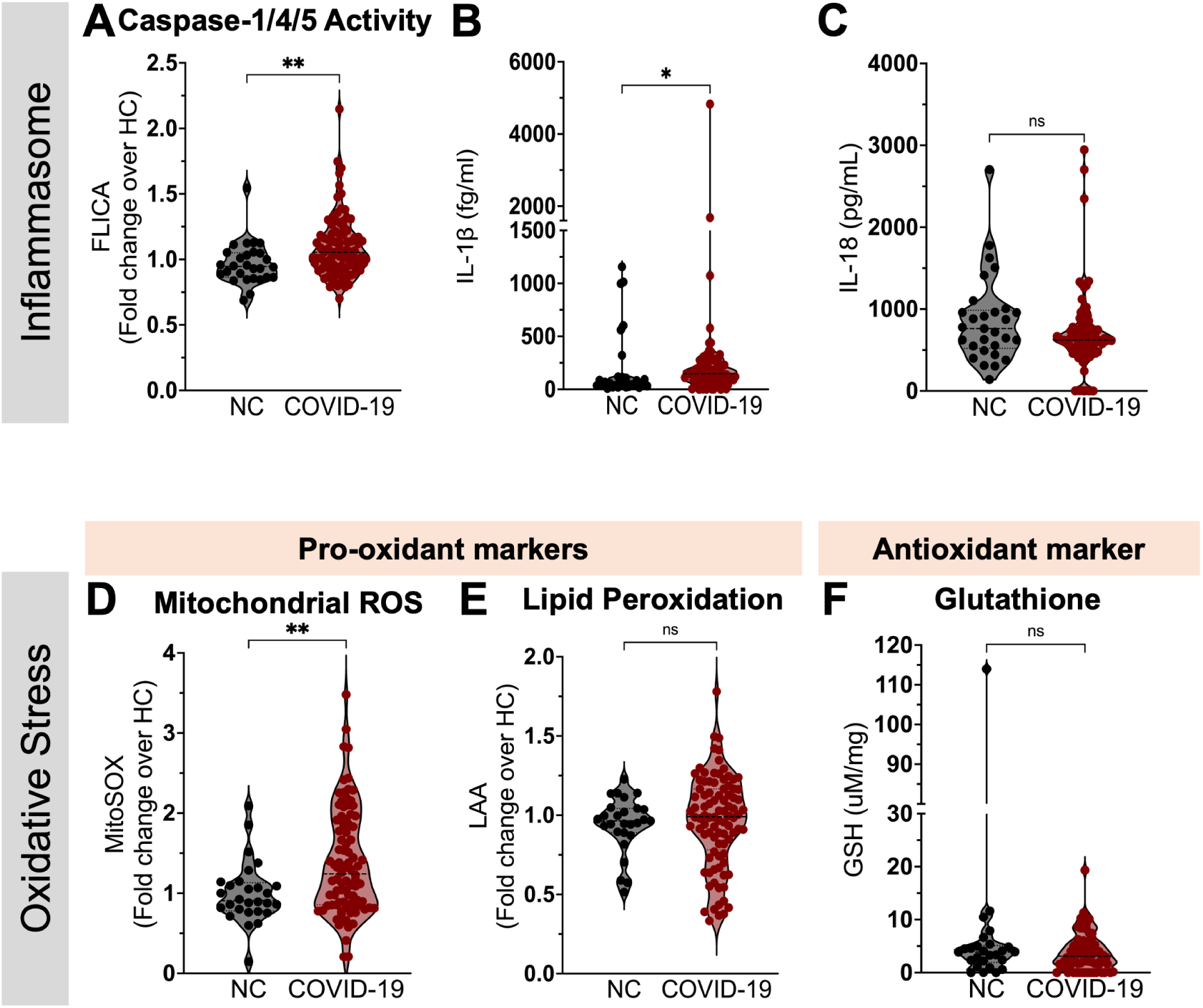
Measures of inflammasome activity and oxidative stress in monocytes from participants with and without prior COVID-19. (**A**) Geometric mean fluorescence intensity (gMFI) of FLICA (fluorochrome inhibitor of caspase-1/4/5) within circulating monocytes from SARS-CoV-2-naïve controls (NC, n=28) and COVID-19 participants (n=92). Plasma levels of IL-1β (**B**) and IL-18 (**C**) in NC (n=29) and COVID-19 participants (n=87). Levels of mitochondrial superoxide (MitoSOX) **(D)** and Lipid peroxidation (95) **(E)** in monocytes from NC (n=28) and COVID-19 (n=92). (**F**) Intracellular levels of glutathione (GSH) were measured in PBMC lysates from NC (n=26) and COVID-19 participants (n=69). gMFIdata are expressed as median of the fold change (gMFI of each sample over the gMFI mean of the NC group). Bars represent median ± quartiles. Statistical analysis was performed using a two-sided Mann-Whitney test (*, *P* < 0.05, **, *P* < 0.005).

To investigate the oxidative stress pathway in circulating monocytes from study participants, we first evaluated the generation of the primary reactive oxygen species (ROS), mitochondrial superoxide (MitoSOX). In comparison to naïve controls, we found increased intracellular levels of MitoSOX in monocytes from individuals with prior COVID-19 (Figure 2D). As a result of elevated levels of mitochondrial superoxide, an excessive production of highly reactive hydroxyl radicals occurs, which in turn react to membranes enriched in polyunsaturated fatty acids, leading to the generation of lipid hydroperoxides. This process of lipid peroxidation is limited by the use of intracellular glutathione (GSH) by glutathione peroxidase 4 (*28*). Interestingly, opposing to our previous findings in people with acute COVID-19(*8*), we failed to detect an increase of lipid peroxidation by monocytes (Figure 2E) or a defect in intracellular glutathione generation (Figure 2F) in PBMCs from participants with prior COVID-19 as compared to NC. These findings suggest that monocytes do not accumulate lipid ROS, despite persistently higher levels of MitoSOX generation following SARS-CoV-2 infection.

Compared to naïve controls, both RC and LC participants had higher caspase-1/4/5 activity (Figure 3A) and plasma IL-1β levels (Figure 3B). However, those with LC did not differ from RC participants (Figure 3A and 3B). Furthermore, no differences were found in IL-18 levels among all groups tested (Figure 3C). In contrast, MitoSOX levels were elevated only in the group with LC symptoms when compared to NC (Figure 3D), while no differences were found in intracellular levels of lipid peroxidation and glutathione amongst the groups (Figure 3E and 3F, respectively). Besides its regulatory function on cell cycle, the p16^INK4a^ molecule has also been reported to control intracellular ROS production, as loss of p16^INK4a^ results in aberrant mitochondrial mass along with elevated mitochondrial membrane potential (Δψm) and ROS (*29*). Accordingly, we found a significant association between p16^INK4a^ intracellular levels and MitoSOX measurements within participants presenting with LC symptoms (Figure 3G and 3H), suggesting downmodulation of p16^INK4a^ in monocytes may contribute to their dysregulated high ROS production in LC.

**Figure 3.**
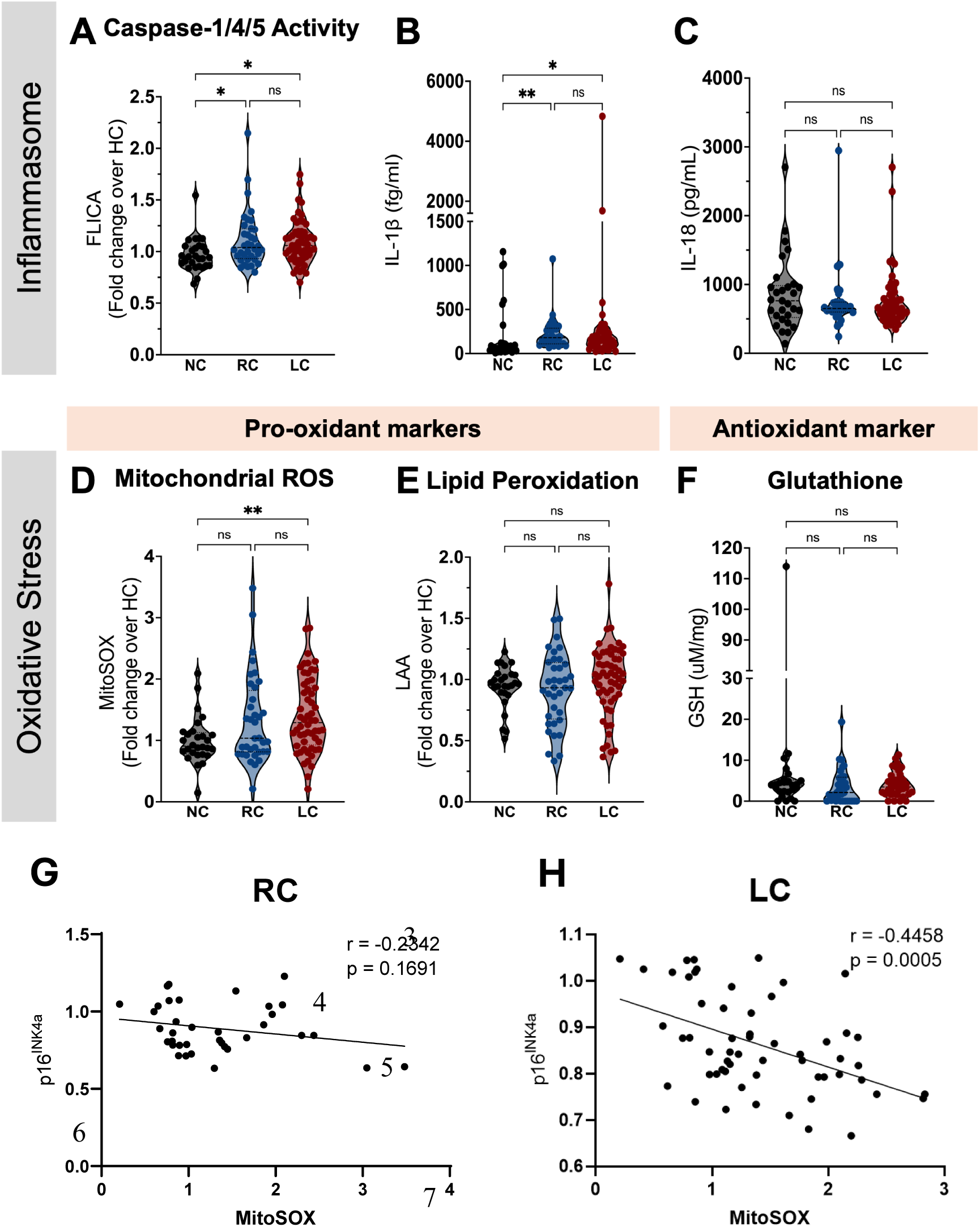
Inflammasome and oxidative stress measurements post-COVID-19, stratified by the presence of symptoms. (**A**) Geometric mean fluorescence intensity (gMFI) of FLICA (fluorochrome inhibitor of caspase-1/4/5) within circulating monocytes from SARS-CoV-2-naive controls (NC, n=28), COVID-19 recovered individuals (RC, n=36) and participants presenting with long COVID symptoms (LC, n=56). Plasma levels of IL-1β (**B**) and IL-18 (**C**) in NC (n=29), RC (n=29) and LC (n=49). Levels of mitochondrial superoxide (MitoSOX) **(D)** and Lipid peroxidation (95) **(E)** in monocytes from NC, RC and LC. (**F**) Intracellular levels of glutathione (GSH) were measured in PBMC lysates from NC (n=25), RC (n=32) and LC (n=37). gMFIdata are expressed as median of the fold change (gMFI of each sample over the gMFI mean of the NC group). Bars represent median ± quartiles. Statistical analysis was performed using a Kruskal-Wallis test (*, *P* < 0.05, **, *P* < 0.005). Spearman correlations between intracellular levels of p16^INK4a^ and MitoSOX measured in total circulating monocytes from COVID-19 participants stratified into RC **(G)** or LC **(H)**.

In summary, these findings demonstrate an enrichment of immature monocytes (HLA-DR^lo^/p16^INK4alo^) and sustained inflammasome activity and oxidative stress in individuals during the post-acute phase of COVID-19, with a clear association between low p16^INK4a^ and dysregulated ROS production in those experiencing LC.

### Subhead 3: Evidence of systemic chronic inflammation, oxidative stress and senescence following COVID-19

To further assess the role of SARS-CoV-2 infection in immune disruption, we evaluated levels of plasma biomarkers from study participants. Consistent with previous reports (*30, 31*) and our cellular findings, we found evidence of systemic monocyte activation following COVID-19 with elevated pro-inflammatory mediators including the monocyte activation markers sCD14 (Figure 4A) and sCD163 (Figure 4B) in those with prior COVID-19 when compared to naïve controls. The type I IFN signaling chemokine, IP-10, was also significantly elevated in individuals with prior COVID-19 (Figure 4C). Furthermore, irisin, an indicator of increased metabolic rate in monocytes and marker of oxidative stress (*32*), was significantly elevated in those with prior COVID-19 (Figure 4D) as was the secreted protein acidic and rich in cysteine, (Figure 4E, SPARC) which has been shown to convert macrophages to a pro-inflammatory phenotype and to dampen mitochondrial respiration (*33*). In contrast, soluble urokinase plasminogen activator receptor (suPAR), a biomarker of systemic chronic inflammation (*34*), was not altered in individuals with prior COVID-19 compared to naïve controls (Figure 4F).

**Figure 4.**
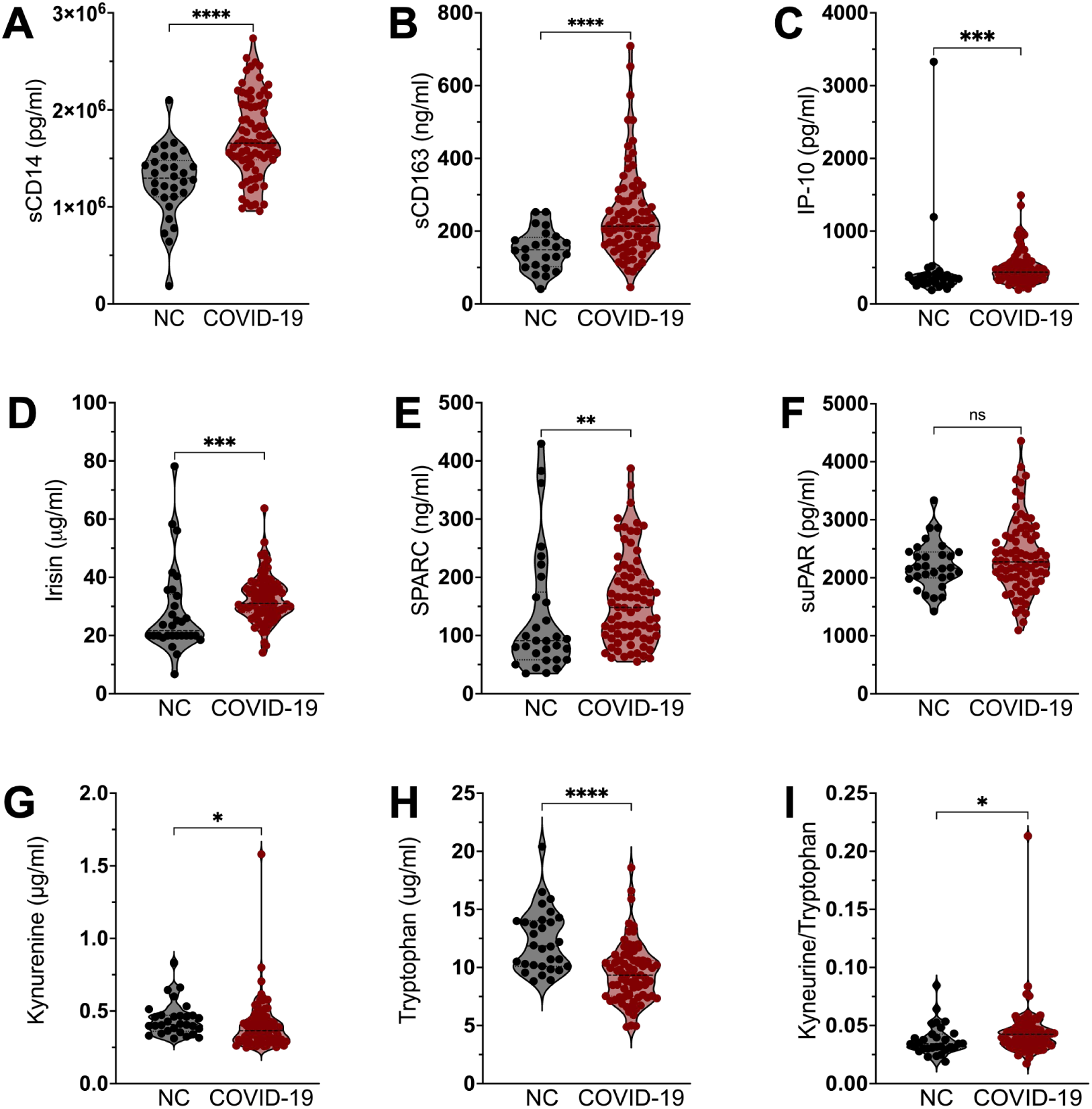
Plasma biomarkers compared between COVID-19 participants and no prior COVID controls (NC). Plasma concentrations of (**A**) sCD14, (**B**) sCD163, (**C**) IP-10, (**D**) Irisin, (**E**) SPARC, (**F**) suPAR, (**G**) Kynurenine, and (**H**) Tryptophan in SARS-CoV-2-naive controls (NC) compared to COVID-19 patients. (**I**) Calculated Kynurenine/Tryptophan ratio in NC compared to COVID-19 patients. Bars represent median ± quartiles. Statistical analysis was performed using a two-sided Mann-Whitney test (*, *P* < 0.05, **, *P* < 0.005, ***, *P* < 0.0005, **** *P* < 0.0001).

It has been previously reported that decreased circulating levels of tryptophan (Try) and its metabolite serotonin, along with increased levels of its catabolites, kynurenine (Kyn) and kynurenic acid (Kyna), associate with severe COVID-19 and poor long-term outcomes (*35, 36*). Here we observed increased Kyn and remarkably decreased Try plasma levels in participants with prior COVID-19 when compared to NC (Figure 4H and 4I), along with significantly higher Kyn/Try ratio, (Figure 4J), corroborating that an altered Try pathway persists following SARS-CoV-2 infection.

Alterations in the plasma biomarkers of inflammation investigated above remained significant when individuals with prior COVID-19 were divided between RC and those who had LC (Figure 5). However, no differences in sCD14, sCD163, IP-10, Irisin or SPARC were noted between RC and LC individuals (Figure 5A - E). In contrast, we observed a significantly higher suPAR level in LC individuals compared to RC (Figure 5F). Furthermore, individuals with LC symptoms showed a more pronounced dysregulation in the Kyn and Try levels (Figure 5H and 5I), as previously shown for our LC study participants (*37*) and other cohorts of LC patients (*37, 38*).

**Figure 5.**
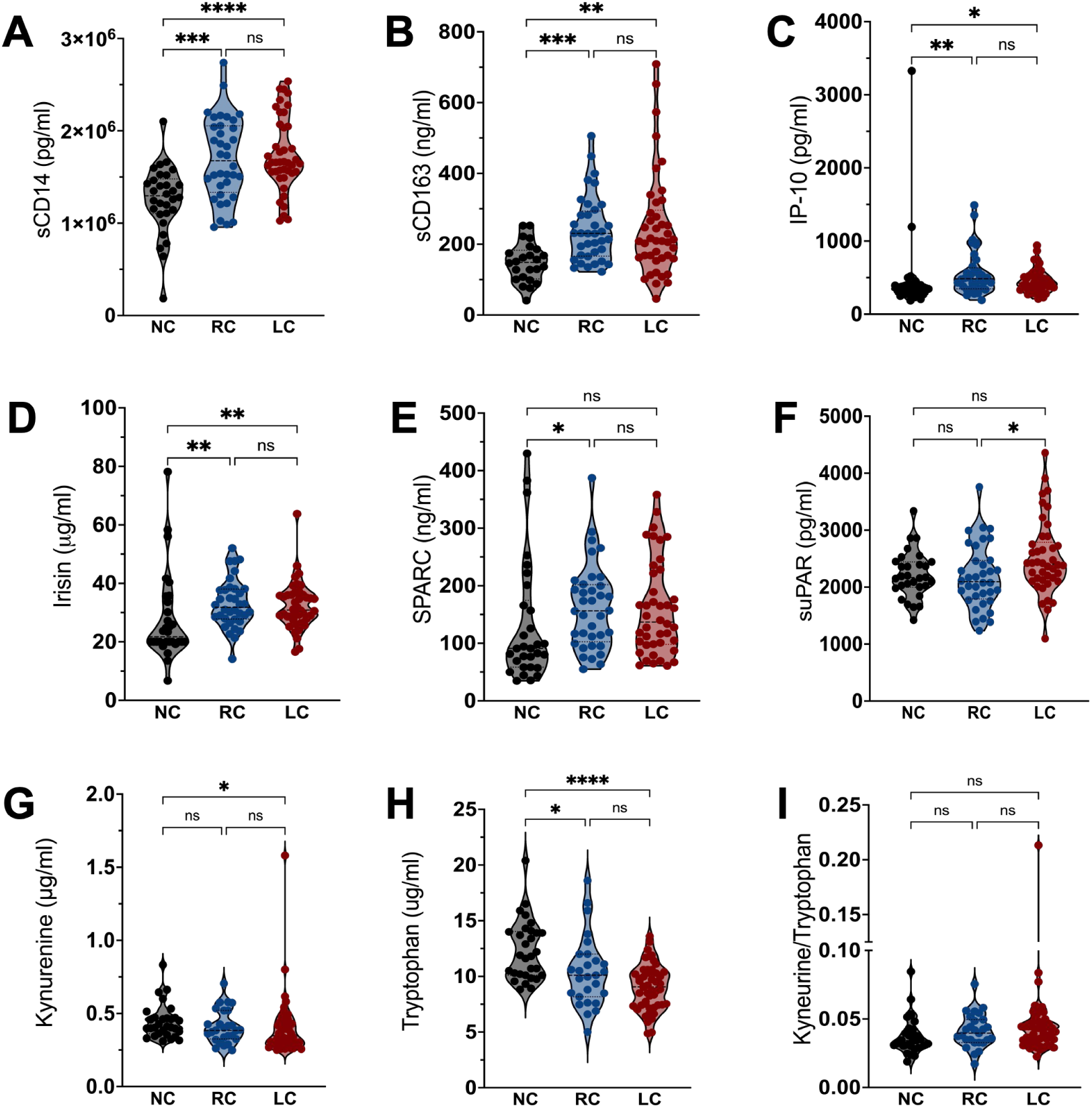
Plasma biomarker concentrations in no prior COVID (NC), recovered (RC), and Long COVID (LC) participants. Plasma concentrations of (**A**) sCD14, (**B**) sCD163, (**C**) IP-10, (**D**) Irisin, (**E**) SPARC, (**F**) suPAR, (**G**) Kynurenine, and (**H**) Tryptophan in SARS-CoV-2-naive controls (NC), recovered COVID-19 (RC), and Long COVID (LC) patients. (**I**) Calculated Kynurenine/Tryptophan ratio comparison by group. Bars represent median ± quartiles. Statistical analysis was performed using a Kruskal-Wallis test (*, *P* < 0.05, **, *P* < 0.005, ***, *P* < 0.0005, **** *P* < 0.0001).

A broad panel of lipid mediators of inflammation was also evaluated. A heatmap with unsupervised clustering analysis identified naïve controls from those with prior COVID-19; those with prior COVID-19 generally clustered together regardless of LC status, and formed two unique clusters flanking the NC group (Figure 6). One cluster had relative increases in several lipid mediators which are known to be associated with increased oxidative stress and which can propagate myeloid inflammation, however these differences were less pronounced in the other cluster. Individual bar plots highlight that compared to naïve controls, classic inflammatory lipid mediators (arachidonic acid [AA], 5-hydroxyeicosatetraenoic acid [5-HETE], 12-HETE, 11,12-dihydroxyeicosatreinoic acid [11,12-DiHETrE]) were elevated in individuals with prior COVID-19, whereas the levels of lipid mediators with anti-inflammatory properties (arachidonoyl ethanolamide [AEA], docosahexaenoyl ethanolamide [DHEA]) were decreased (Supplemental Figure 2).

**Figure 6.**
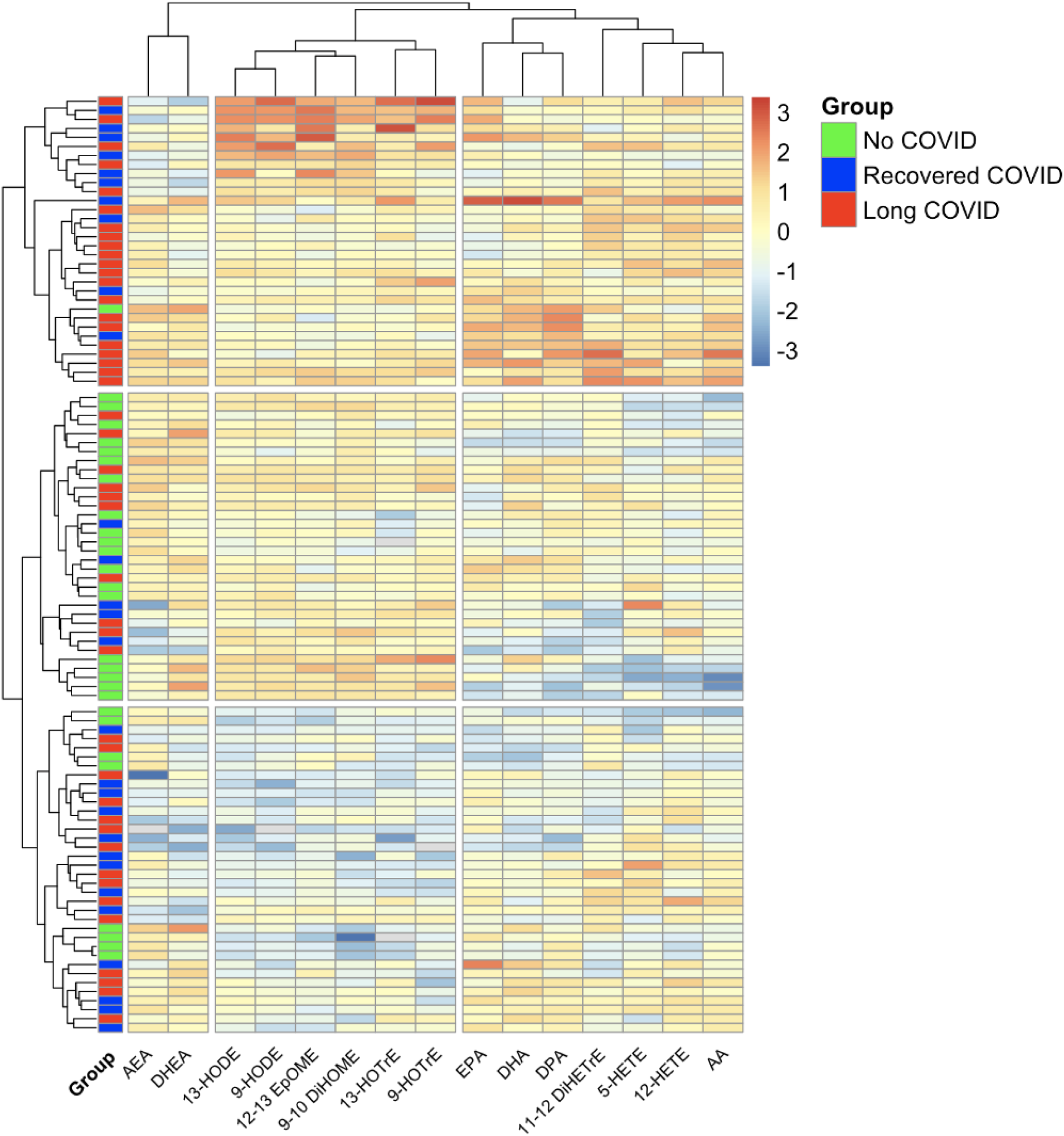
Lipid mediators of inflammation displayed using a heatmap with unsupervised clustering. Individuals are represented by colored boxes and unsupervised clustering analysis highlights those with no prior COVID-19 (NC) primarily cluster with two unique clusters on each end representing prior COVID-19 individuals. There is no unique clustering between individuals recovered from COVID-19 (RC) vs those with long COVID (LC). Heatmap generated using R (version 4.3.2) with the pheatmap package. HODE, hydroxyoctadecadenoic acid; EpOME, epoxyoctadecamonoenoic acid; DiHOME, dihydroxyoctadecenoic acid; HOTrE, hydroxyoctadecatrienoic acid, DHA, docosahexaenoic acid; DPA, docosapentaenoic acid; HEPE, hydroxyeicosapentaenoic acid; EPA, eicosapentaenoic acid; DiHETrE, dihydroxyeicosatreinoic acid; HETE, hydroxyeicosatetraenoic acid; AA, arachidonic acid.

### Subhead 4: LC associates with chronic inflammation, sustained T-cell activation and CD8 T-cell senescence

We next compared immune cell profiling in peripheral blood samples from participants with prior COVID-19 to naïve controls using high-dimensional flow cytometry. We identified 25 clusters representing T-cells, B-cells, NK cells, innate lymphoid cells (ILCs), plasmacytoid dendritic cells (pDCs), and monocytes (Figure 7A). Clustering analysis identified two distinct subsets of early effector memory (EM) CD4 T-cells, CD45RA^+^ terminal effector (RATE) CD8 T-cells, EM CD8 T-cells, γγγγT-cells, and mature NK cells that would not have been distinguished through manual gating. Of the 25 metaclusters (MC), we identified 3 distinct MC which significantly differentiated prior COVID-19 from NC (Figure 7B). We observed a decrease in the frequency of pDC (Figure 7C) and naïve CD4 T-cells (Figure 7D) and elevated EM CD8 T-cells in those with prior COVID-19 compared to naïve controls (Figure 7E).

**Figure 7.**
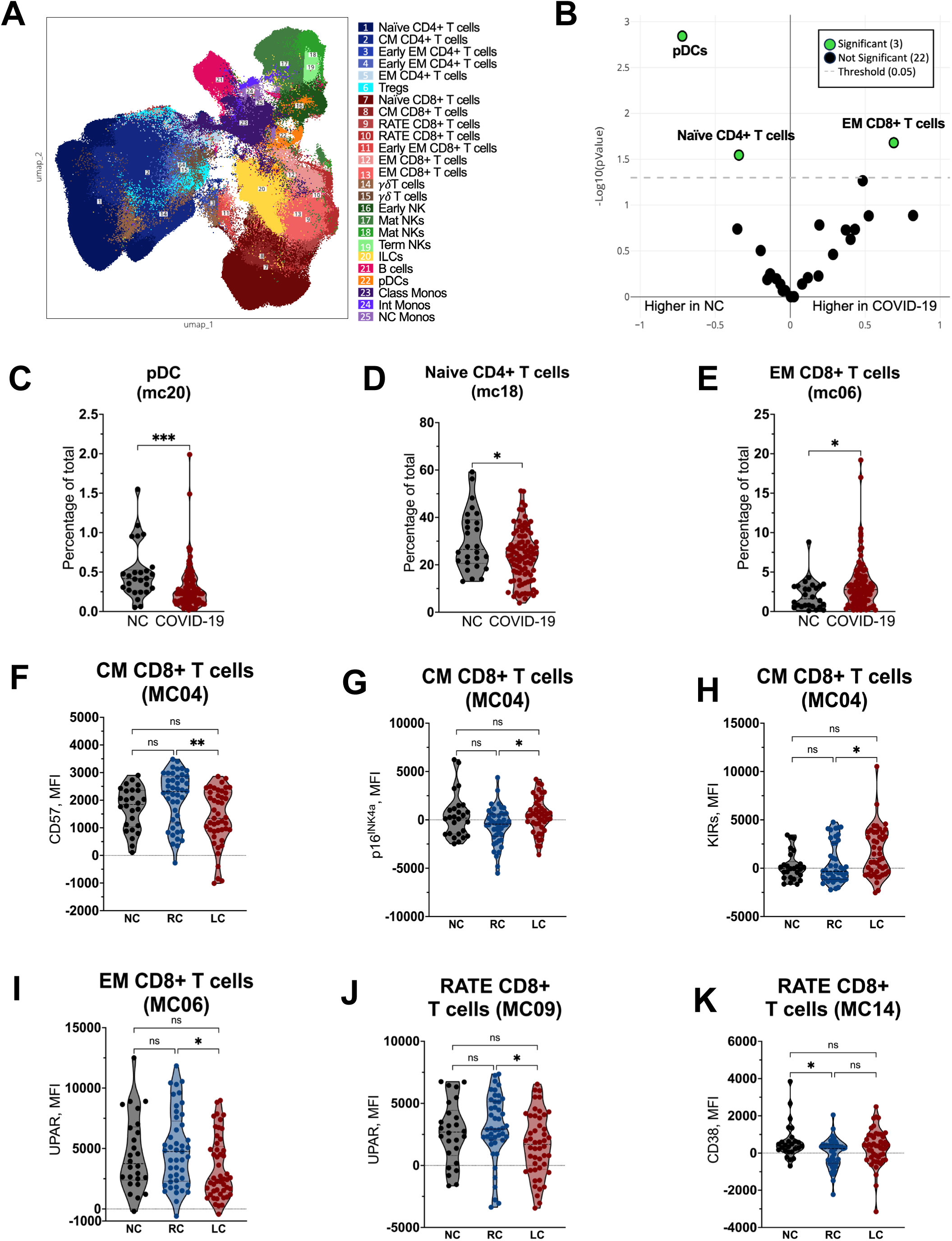
Immune phenotyping of PBMCs following COVID-19 infection. (**A**) UMAP dimensionality reduction embedding of peripheral blood mononuclear cells (PBMCs) from the entire dataset colored by MC. (**B**) Volcano plot showing differentially expressed MC between SARS-CoV-2-naive (NC) and COVID-19 patients. Log2 fold change (FC) is represented on the x-axis and *P value* is represented on the y-axis (*P* < 0.05 considered significant). Frequency of (**C**) pDCs (MC20), (**D**) naïve CD4 T cells (MC18), and (**E**) effector memory (EM) CD8 T cells (MC06) in SARS-CoV-2-naive controls (NC) and COVID-19 patients. Analysis was performed using a two-sided Mann-Whitney test. *, *P* < 0.05, ***, *P* < 0.0005. (**F**) CD57, (**G**) p16^INK4a^, (**H**) killer immunoglobulin receptors (KIR) in central memory (42) CD8 T cells (MC04), (**I**) UPAR in effector memory (EM) CD8 T-cells (MC06), and (**J**) UPAR and (**K**) CD38 in CD45RA+ terminal effector (RATE) CD8 T-cells in SARS-CoV-2-naïve controls (NC), recovered COVID-19 (RC), and Long COVID (LC) patients. Bars represent median ± quartiles. Statistical analysis was performed using a Kruskal-Wallis test (*, *P* < 0.05, **, *P* < 0.005).

We next compared expression of markers related to activation, exhaustion, and senescence in identified MC between the prior COVID-19 and naïve control groups. In addition to pDCs being less abundant in those with prior COVID-19, we found significantly lower expression of CD38 and higher PD-1 expression in the pDCs in this group compared to naïve controls (Supplemental Figure 3A and 3B). This indicates that circulating pDCs from individuals with prior COVID-19 were more activated than those in naïve controls.

In CD4 T-cells, we observed higher levels of activation, measured through HLA-DR in the less differentiated central memory CD4 T-cells (Supplemental Figure 3C), but lower levels of Ki67 in further differentiated early EM and EM CD4 T-cells (Supplemental Figure 3D and 3E). The senescence marker p16^INK4a^ was significantly lower in effector memory CD4 T-cells of those in the group with prior COVID-19 compared to naïve controls (Supplemental Figure 3F). Finally, we saw elevated HLA-DR in RATE CD8 T-cells of those with prior COVID-19 compared to naïve controls (Supplemental Figure 3G). Taken together, these data indicate evidence of persistent perturbations in the frequency and activation of peripheral immune cells in the post-acute phase of COVID-19, months after acute infection.

We further compared the immune profile of recovered and LC participants. The most profound differences between these groups were found in the CD8 T-cell subset (Figures 7F – K). In CM CD8 T-cells, we observed an elevation in p16^INK4a^ and killer-cell immunoglobulin-like receptors (KIRs) and lower CD57 expression in those with LC compared to those who fully recovered. In both EM and RATE CD8 T-cells, UPAR expression was significantly lower in those with LC compared to those who recovered. Finally, CD38 expression was significantly lower in fully recovered individuals compared to naïve controls, but no differences were observed between LC and these other groups.

We next selected a representative subset of matched individuals with Long COVID versus full recovered (n=7 per group) to perform single cell RNA-sequencing analysis (scRNA seq). Gene set enrichment analysis (GSEA) identified distinct biological processes enriched in the T-cell compartment of LC indicating sustained activation. In CD4 T-cells, 12 pathways were significantly enriched in LC compared to RC (Figure 8A), including gene sets related to chromatin organization and immune response; specifically, multiple pathways related to immune activation including complement, humoral immunity, and signaling pathway activation. GSEA identified 21 pathways enriched in CD8 T-cells from LC individuals (Figure 8B), including pathways related to adhesion and chemotaxis, further supporting sustained activation in this subset. Apoptotic cell clearance was also significantly enriched in CD8 T-cells of those with LC, which may reflect an ongoing response to cellular turnover and tissue damage. Interestingly, within the monocyte compartment, numerous pathways were downregulated in LC compared to full recovery, including endocytosis, response to external stimuli, regulation of inflammatory responses, cytokine-mediated signaling pathway and myeloid activation (Figure 8C). This confirms a profoundly altered myeloid profile in LC.

**Figure 8.**
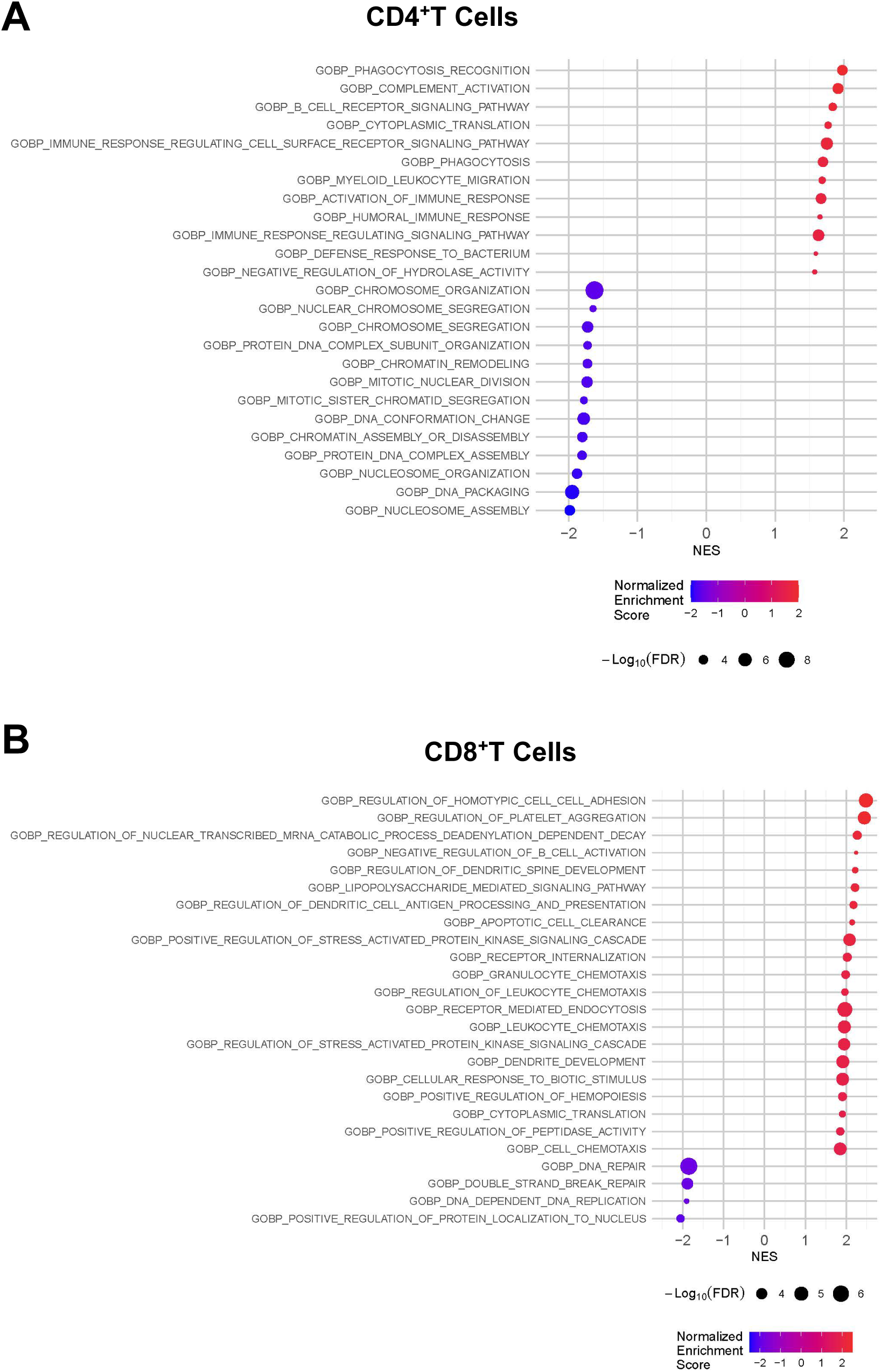
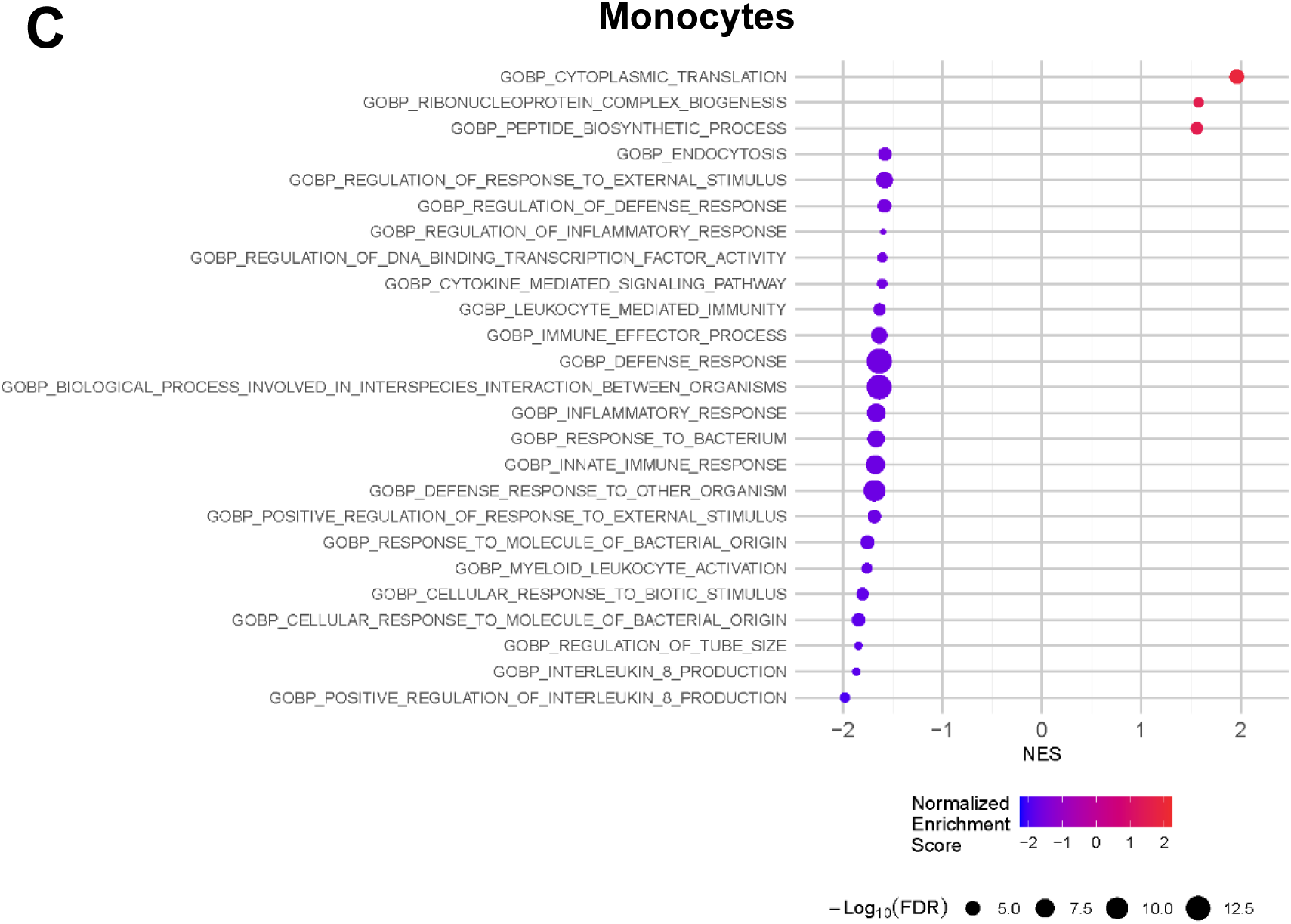
Transcriptional profile of study participants reveals differences from LC participants when compared to recovered (RC) group. Single cell suspensions were obtained from PBMCs derived from recovered (n=7) and Long-COVID (n=7). Enriched pathways identified by GSEA when comparing LC and recovered groups in CD4^+^ **(A)**, CD8^+^ T cells **(B)** and monocytes **(C)**. Dot sizes represent FDR values and positive Normalized Enrichment Scores depict higher expression in LC.

Finally, an exploratory analysis using Elastinet regression models incorporating variables that demonstrated significant differences between groups was performed, on biomarker and immunophenotyping datasets, to identify which markers most strongly differentiated those with prior COVID-19 vs NC. For the immunophenotyping data, five markers (percent classical monocytes, classical monocyte expressing CD163, and patrolling monocytes, total monocyte p16^INK4a^ expression, and total monocyte HLA-DR expression) accounted for over 60% of the variability within the cohort when incorporated into a principal component analysis (Figure 9A). A separate model that included soluble biomarkers identified 7 biomarkers (AA, sCD14, 5-HETE, 12-HETE, Try, DHEA, AEA) accounted for approximately 60% of the variability (Figure 9B). Elastinet modeling was repeated comparing those with the fully recovered vs LC cohort although differences between these two groups were much less prominent. Using the immunophenotyping data, four variables (percent classical monocytes, CD57 expression on Tregs and CM CD8 T-cells, and CM CD8 p16^INK4a^) accounted for 71% of the variability with low separation between groups by PCA. Five biomarkers (AA, AEA, Try, IP10, and 11-12-DiHETrE) provided the greatest differentiation between the recovered COVID-19 and LC subgroups, although a PCA constructed using these variables still showed overlap between these groups highlighting that the differences, albeit present, were subtle (Figure 9C).

**Figure 9.**
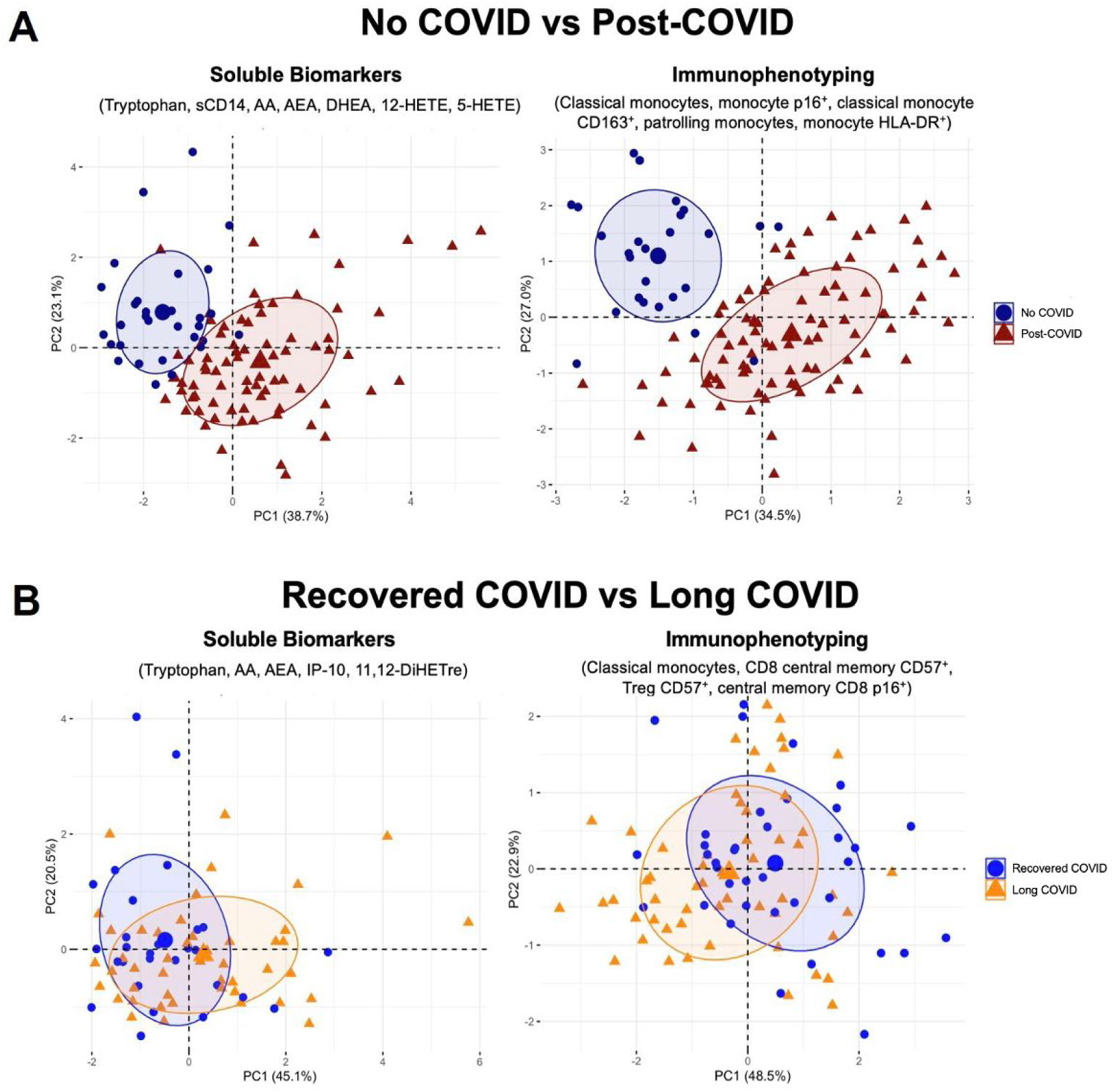
Principal component analysis highlights unique immune profiles in No COVID vs Post-COVID groups. Principle component analysis (PCA) was performed incorporating the most differentiating soluble biomarkers and immunophenotyping markers selected by Elastinet regression models. PCAs were generated in R (version 4.3.2) using the FactoMineR and factoextra packages. **(A)** PCA comparing No COVID (NC) to post-COVID patients. (**B**) PCA comparing recovered COVID-19 (RC) and Long COVID (LC) patients. AA, arachidonic acid; AEA, arachidonoylethanolamide; DHEA, dehydroepiandrosterone; 5,12-HETE, 5,12-hydroxyeicosatetraenoic acids; IP-10, interferon gamma-induced protein 10; 11,12-DiHETre, dihydroxyeicosatrienoic acids.

### Subhead 5: Changes in DNA methylation patterns occur in those with prior COVID-19

Previous reports have implicated advanced epigenetic aging following COVID-19 (*15, 16*), therefore we decided to further evaluate DNA methylation (DNAm) in blood samples from a subset of 88 total patients (31 NC, 30 recovered COVID-19, and 27 LC). Epigenetic age was estimated using the Horvath clock and correlated strongly with chronological age in each group (Supplemental Figure 4A and 4B). No significant differences were observed in the Horvath age of COVID-19 patients compared to NC (Supplemental Figure 4C) or between RC and LC (Supplemental Figure 4D) in our cohort.

Significant differences in the gene methylation patterns, however, were found between the group with prior COVID-19 and naïve controls (Figure 10A). Figure 10B and Supplemental Table 2 highlight the most differentially methylated CpGs and their respective genes. Among them, several genes associated with various intracellular processes were found to be highly methylated in those with prior COVID-19 as compared to naïve controls (Figure 10B). Those include genes involved in cellular metabolism such as *PCED1A* (*39*)*, GLB1L* (*40*), and *IGFBP* (*41*) as well as anti-inflammatory immune responses (*ZC3H8*) (*42*). The SLC12A family of genes which play a functional role in the pathophysiology of hypertension and cardiovascular disease (*43*) and VPS16, which encodes a protein involved in vesicle trafficking, are also of particular interest, since changes in these genes have been associated with neurodevelopmental diseases and dementia (*44–46*).

**Figure 10.**
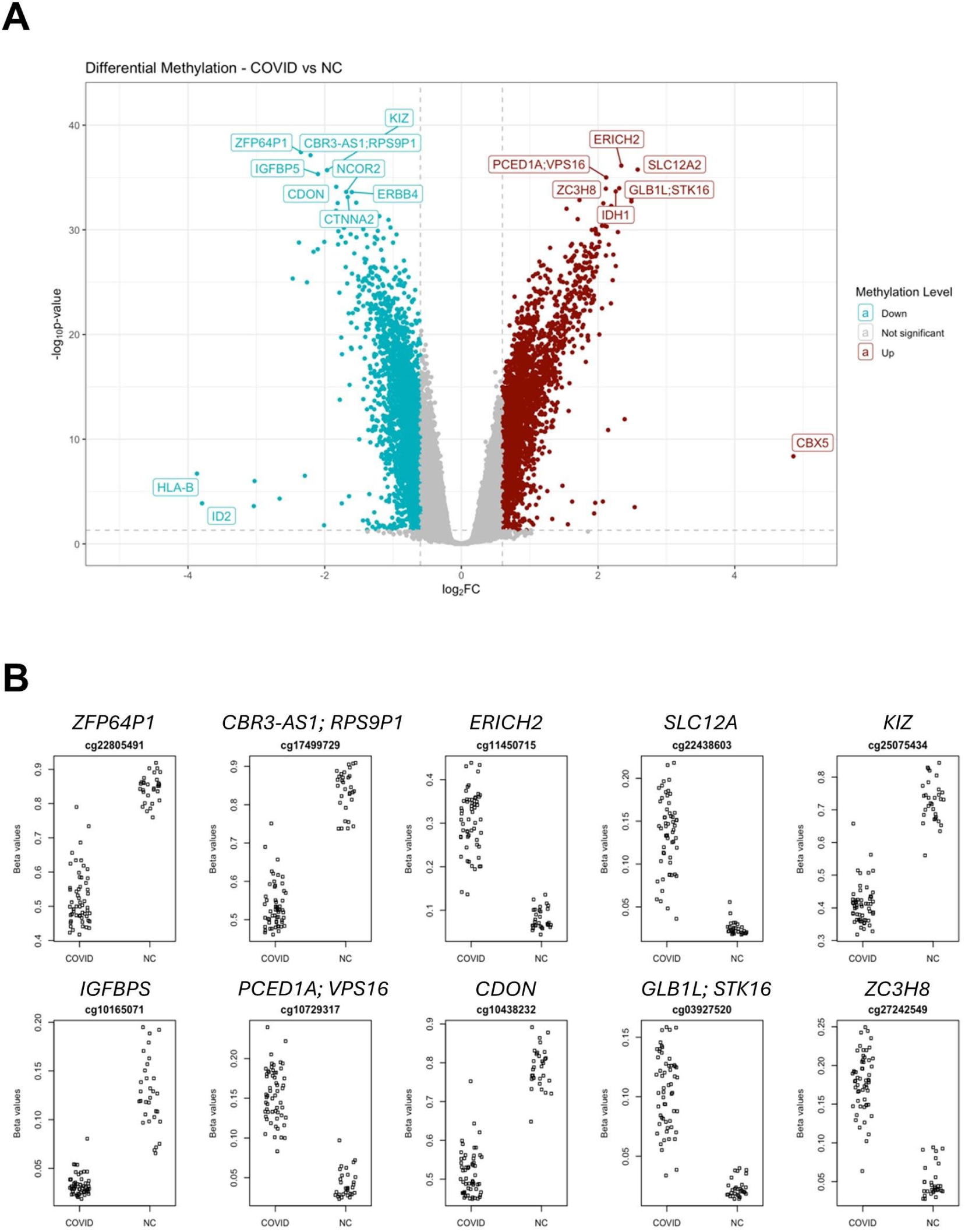
Differential methylation analysis between COVID and NC. **(A)** Volcano Plot displaying the differentially methylated CpGs between prior COVID and NC. Y-axis shows significance represented by −log10(p-value) and X-axis shows relative difference in methylation levels, represented by log2(fold change). CpGs are colored by COVID methylation level, relative to the NC group. The thresholds for “not significant” are: p-value > 0.05 or abs(log2FC) < 0.6. The top CpGs are labeled with the gene symbol. **(B)** Graphs show the top ten differentially methylated CpGs between COVID and NC (y-axis = CpG beta values; x-axis = Sample groups). The beta values reflect the methylation level of the labeled CpG. The gene name is labeled above each plot.

When compared to naïve controls, several genes involved in promoting oncogenesis showed decreases in methylation in those with prior COVID-19 (Figure 10B). The Zinc finger protein 64 (ZFP64) is a transcription factor shown to positively regulate the development of multiple cancers, including hepatocellular carcinoma (HCC) and esophageal cancer (*47–49*). ZFP64 activates the promoter of the mixed spectrum leukemia (MLL) oncogene, thus maintaining its high expression level (*50*) as well as the promoter of Gal-1, PD-1, and CTLA-4 genes, thereby rendering cancer cells resistant to immunosuppression (*49–51*). Likewise, the CBR3 Antisense RNA 1 (CBR3-AS1) has been characterized as an oncogenic long non-coding RNA in several cancers (*52–55*), and also participates in chronic inflammatory and autoimmune diseases such as ulcerative colitis (*56*) and rheumatoid arthritis (*57*). Decreased methylation, and therefore increased expression of tumorigenic genes in those with prior COVID-19 when compared to naïve controls, could potentially indicate -especially if persistent- an increased risk of malignancy development months following SARS-CoV-2 infection.

Gene Ontology (GO) pathway analysis further highlights that differentially methylated genes between the groups primarily comprise those involved with several cellular components (CC), including intracellular and nuclear organelles and protein-containing complexes as well as those involved in distinct biological processes (BP), such as metabolism, biosynthesis, and chromatin organization (Supplemental Table 3).

## DISCUSSION

Although the profound inflammatory response in the acute phase of SARS-CoV-2 infection has been well characterized (*8, 22, 58, 59*), the role of SARS-CoV-2 in causing sustained immunologic changes in the post-acute phase, including in the setting of Long COVID, is less well-understood. Here, we demonstrate extensive alterations approximately 4 months following an initial SARS-CoV-2 infection. Post-acute metabolic derangements included markedly decreased Try levels, high levels of pro-inflammatory lipid mediators and epigenetic changes in cellular metabolism-related genes. Immunologic perturbations were marked by systemic myeloid inflammatory responses, persistent activation of peripheral immune cells, sustained inflammasome and oxidative stress activity, and increases in circulating inflammatory markers. We found Long COVID to be associated with sUPAR and immune senescence markers in CD8 T-cells, as well as dysregulated monocyte and T-cell populations. Finally, we observed a differential methylation pattern in participants with prior-COVID-19, suggesting cellular homeostatic imbalance as well as a potential increased risk for malignancies. Overall, our findings demonstrate the long-term metabolic and immunologic consequences that can follow an acute viral infection, and describe a post-acute pathophysiologic substrate characterized by chronic innate immune activation, metabolic distress, and T-cell senescence that may contribute to LC and other post-acute sequelae as well as possible higher susceptibility to new infections.

We identified prominent post-acute metabolic dysfunction that may contribute to adverse outcomes from COVID-19. For example, our analysis using Elastinet regression models identified that dysregulation in lipid mediators of inflammation provided the greatest differentiation between the Long COVID and fully recovered groups. AA and 11-12-DiHETrE, found to be increased post-COVID, play a crucial role in inflammatory and allergic reactions (*60*), and have been implicated in the pathogenesis of many chronic inflammatory diseases (*61, 62*). In contrast, AEA and DHEA, which are derived from AA and omega-3 fatty acids [ω-3 FAs], respectively, and associated with beneficial cardiovascular, neurological and anti-inflammatory effects (*63*), were lower in those with prior COVID-19 compared to naïve controls. Collectively these observations suggest that treatments targeting lipid pathways, such as omega-3 supplementation or agents like metformin which have been shown to prevent LC (*64*), could provide benefit as they have been found to positively impact AA metabolism (*65*).

Notably, low levels of ω-3 FAs have also been associated with pathophysiology of myalgic encephalomyelitis/chronic fatigue syndrome (ME/CFS) (*66*), a related infection-associated chronic condition with immunopathology involving mitochondrial, metabolic and immune dysfunction (*67, 68*). Tryptophan metabolism has also been implicated in ME/CFS pathology (*69*). Reduced circulating Try leads to depletion of serotonin levels, potentially contributing to neurologic and cognitive symptoms observed in ME/CFS and LC (*37*). Since LC can comprise a greater variety of symptoms than ME/CFS, future studies should evaluate the contribution of altered metabolic profiles in additional LC cohorts presenting specifically with ME/CFS phenotype.

Our immunologic findings support the hypothesis that persistent viral antigens or tissue damage may promote lingering innate immune responses and systemic low-grade inflammation following SARS-CoV-2 infection, although it remains unclear to what extent this is mechanistically tied to LC symptoms. A shift towards an immature myeloid blood profile is commonly seen in viral infections, including acute SARS-CoV-2 (*70–73*). The presence of a similar pattern months into the post-acute phase may indicate prolonged myelopoiesis (PM), a characteristic of chronic inflammatory conditions like cancer, chronic infections, and autoimmune diseases. IL-1β, which we found to be increased in those with LC, has been reported to promote PM through sustained proliferation and differentiation of hematopoietic stem cells (*74*), driven by persistent antigen or tissue injury. Recently, the presence of SARS-CoV-2 antigens has been reported in the blood of some individuals for up to 14 months post-COVID (*75–77*); other studies have identified a higher occurrence of serologic markers suggestive of EBV reactivation in patients with LC (*78, 79*). Despite these findings, no differences in inflammatory mediators or the proportion of immature monocytes were observed between recovered and LC participants in our cohort. Still, our findings provide further rationale for clinical trials targeting SARS-CoV-2 persistence or the reactivation of latent herpesviruses in LC.

Beyond viral persistence, chronic physiologic stress and inflammation can be driven by prolonged tissue injury including gut disruption with potential bacterial lipopolysaccharide (LPS) or fungal translocation. Persistent gut damage caused by SARS-CoV-2 infection was shown to induce long-term dysregulation of Try absorption from the intestine (*37*), thus potentially explaining the drastic decrease in plasma levels of Try that we observed in the LC group. Additionally, AA has been shown to alter the gut microbiota, increase pro-inflammatory cytokines, and induce moderate neuroinflammation in mice (*80*). Regardless of microbial translocation, chronic tissue injury, including persistent endothelial dysfunction, can also release endogenous damage signals from dying/compromised cells, thus leading to systemic oxidative stress and inflammasome activation (*81*). Therefore, collectively, our findings support treatments targeting oxidative stress (*82*) and inflammasome activation in LC, as well as the use of immune modulators, that act upstream of inflammasome and other inflammatory pathways and may impact emerging myelopoiesis (*83*), to help mitigate broad inflammatory responses post-COVID-19. Herpesvirus reactivation or prothrombotic tendency, however, may be important limitations. Chronic T-cell activation not only contributes to inflammation but also results in T-cell exhaustion. Our data suggest that an unbalanced innate and adaptive immune recovery following SARS-CoV-2 infection leads to a senescent phenotype with impaired CD8 function, potentially contributing to LC symptoms. Recent findings from our cohort demonstrated that individuals with LC exhibit exhausted SARS-CoV-2-specific CD8 T-cells (*13*). In the present study, stratification of individuals with prior COVID-19 revealed a distinct immunological signature within the LC subgroup further validated through single cell analysis. Expression of some markers commonly associated with cellular senescence, such as UPAR and CD57, were significantly lower throughout multiple immune cells including some NK and CD8 T-cell subsets, which may reflect improper activation and proliferation of these effector subsets following antigenic stimulation during acute COVID-19. A similar phenomenon has been reported in chronic HIV infection resulting in less well-differentiated memory cells and poorer killing capacity (*84*). Our single cell sequencing analysis further supports dysregulation and sustained immune activation in T cells during LC, a phenomenon not observed in the monocyte compartment, which inversely, seem to reflect an abundance of immature monocytes with impaired effective responses that may also contribute to decreased immune surveillance.

We also observed evidence of immune senescence in LC. Both P16^INK4a^, a cell cycle regulator specifically recognized as a marker of replicative senescence (*85*), and KIRs, which have also been reported to accumulate on CD8 T-cells during aging (*86*), were significantly elevated in T-cells in those with LC. Additionally, suPAR, a marker of chronic inflammation and immune senescence that has been previously implicated in acute COVID-19 severity (*87*), was the only plasma biomarker that was significantly elevated in LC compared to recovered individuals. Therefore, our findings provide further evidence for the use of immunomodulators like mTOR inhibitors that regulate autophagy and cellular senescence, can influence AA metabolism and are known for anti-tumor activity without reactivation of herpesviruses (*88, 89*) in carefully designed studies aiming to improve LC outcomes.

Despite increased expression of cellular senescence markers, DNAm analysis did not show advanced epigenetic aging in LC as compared to recovered individuals. Significant variation in methylation patterns were observed between prior COVID-19 and NC groups, revealing that genes governing important cellular homeostatic functions, such as cellular ionic balance, endolysosomal fusion, intracellular signaling and metabolic processes, are highly methylated post-COVID. Since dysregulation of these pathways was found to promote inflammatory, metabolic, neurological, and cardiovascular disorders, downmodulation of these genes post-COVID could underly the pathogenesis of some manifestations of LC. Furthermore, genes involved in promoting tumorigenesis were found to be highly methylated in the NC group as compared to the post-COVID group, suggesting dysregulation of their homeostatic silencing post-COVID. Accordingly, the ability of SARS-CoV-2 proteins to interact with and degrade tumor suppressor proteins, including p53, has been previously reported (*90–92*), further supporting the hypothesis that COVID-19, in conjunction with other carcinogenic events, chronic inflammation and stress and decreased immune surveillance, may predispose to cancer development over time (*93*). The relationship between recurrent COVID-19 and cancer risk requires additional comprehensive longitudinal studies.

The strengths of our study include the evaluation of a wide range of immune responses in a cohort of individuals from the first wave of the COVID-19 pandemic, prior to the emergence of major confounders including vaccinations, reinfections, antiviral treatments, and evolving variants. The contemporaneous uninfected control group was matched for key demographic characteristics. Limitations of this study include the lack of sampling during acute infection to ascertain causality, as well as the lack of other post-viral infection comparators to determine if these post-infectious changes are specific to SARS-CoV-2.

Collectively, our data provide comprehensive insights into the pathophysiology and immunologic dysfunction that persists following acute COVID-19. These immune alterations, including prolonged myelopoiesis, low-grade inflammation, and oxidative stress persist for months post-infection potentially amplified by persistent antigenic stimulation. They are associated with severe metabolic disturbances, marked by low tryptophan and high levels of lipid mediators of inflammation. Chronic stress and immune senescence along with modulation of oncogenic pathways may create a harmful cycle of DNA and tissue damage, impairing future immune responses, and potentially increasing susceptibility to recurrent infections and/or malignancies. Post-COVID-19 prolonged immune activation, mitochondrial dysfunction, and metabolic derangements all represent potential therapeutic targets to prevent or improve the long-term outcomes of individuals post-SARS-CoV-2 infection.

## MATERIALS AND METHODS

### Study Design

Detailed methods of the UCSF LIINC study have been reported previously (*20*). Briefly, any adult in the San Francisco Bay Area is eligible to enroll in LIINC ≥ 2 weeks following test-confirmed COVID-19; all individuals in the current analysis had SARS-CoV-2 infection confirmed with nucleic acid amplification testing. Individuals are enrolled regardless of LC status and assessed at baseline, 3-4 months post-COVID, and then approximately every 4 months thereafter. Assessments include a battery of interviewer-administered questionnaires that include demographic characteristics, medical history, SARS-CoV-2 infection and vaccine history, and detailed assessment of COVID-attributed physical symptoms and quality of life, as described previously (*20*).

In accordance with the World Health Organization consensus definition and the 2024 case definition put forth by the National Academies of Sciences, Engineering, and Medicine, we define Long COVID as the presence of at least one COVID-attributed symptom present ≥ 3 months following COVID-19. Importantly, symptoms that pre-date SARS-CoV-2 infection and are not altered are not considered to represent LC. At each visit, peripheral blood is collected and stored as serum, plasma, and cryopreserved peripheral blood mononuclear cells (PBMCs). These specimens were used for the biological analyses described below.

### Plasma biomarkers measurements

Cryopreserved plasma samples from naive controls (NC) and people with prior COVID-19 (RC or LC) were assessed to measure levels of the following inflammatory biomarkers according to the manufacturer’s instructions: interleukin (IL)-1β and IL-18, tumor necrosis factor-α (TNFα), interferon-gamma-induced protein 10 (IP-10), soluble urokinase Plasminogen Activator Receptor (suPAR) and Eotaxin were all quantified using electrochemiluminescence kits from MesoScale (Gaithersburg, MD, USA); sCD14 and secreted protein acidic and rich in cysteine were quantified using enzyme-linked immunoassay (ELISA) kits from R&D Systems (Minneapolis, MN, USA); sCD163 and Irisin were measured with ELISA kits from Aviscera Bioscience (Santa Clara, CA, USA) and Abcam (Cambridge, CB2 0AX, UK), respectively.

### Metabolomics

Non-targeted metabolomics analysis was performed in cryopreserved plasma samples from NC and people with prior COVID-19 (RC or LC), at Metabolon, Inc using previously published methods(*94*). Briefly, samples underwent methanol extraction, and the resulting extract was used for metabolite analysis by ultra-high-performance liquid chromatography/tandem mass spectrometry in both positive and negative ion modes along with gas chromatography/mass spectrometry to maximize compound detection and accuracy. Metabolites were then identified by comparing the spectral signature of sample metabolites to a reference library at Metabolon, Inc. Spectral peaks were used for metabolite identification by a proprietary visualization and interpretation software. Area under the curve for the spectral peaks was used for metabolite quantification. Raw data generated from peak quantification were then normalized to correct for variation in multi-day experiments, where each compound was normalized to a median equal to one.

### Cell culture and treatments

Cryopreserved ficoll-isolated peripheral blood mononuclear cells (PBMCs) from people with prior COVID or persons without prior COVID-19 (NC) were thawed and resuspended in RPMI-1640 media (Corning, NY, USA) supplemented with 10% heat-inactivated human AB serum (Gemini Bio-Products, West Sacramento, USA) and 0.05% benzonase (MilliporeSigma, USA). Cells were rested for 1 hour at 37 °C and 5% CO_2_ and subsequently plated at 10^6^ cells/well in round bottom 96-well plates (Corning Costar^TM^, MilliporeSigma, USA) followed by immune staining.

### Caspase-1/4/5 activation by flow cytometry

PBMCs were incubated with the fluorochrome inhibitor of caspase-1/4/5 (FAM-FLICA, Immunochemistry technologies (ICT), Bloomington, MN) following the manufacturer’s instructions, for 50 min at 37 °C, to allow for binding of FLICA to activated inflammatory caspases. Cells were washed twice with the FLICA kit wash buffer and then incubated with LIVE/DEAD Fixable AQUA Dead Cells Stain (Thermo Fisher, USA) for 15 min at RT, followed by extracellular staining in PBS + 1% BSA with the following fluorochrome-conjugated antibodies for monocyte phenotyping: anti-CD14 BV605 [Clone: M5E2], anti-CD16 PE-Cy7/BV711 [Clone: 3G8], anti-CD3 PE [Clone: HIT3a] and anti-CCR2 PerCP Cy5.5 [Clone: K036C2] are from BioLegend; anti-CD20 e450 [Clone: 2H7], anti-CD19 e450 [Clone: SJ25C1] and anti-CD2 e450 [Clone: RPA-2.10] from eBioscience; anti-CD56 e450 [Clone: B159], anti-HLA-DR APC-Cy7 [Clone: G46-6] and anti-CD66b e450 [Clone:G10F5] are from BD Biosciences. Cells were fixed and permeabilized with Cytofix/Cytoperm (BD Biosciences, USA) overnight at 4°C. Data were acquired on a BD Fortessa flow cytometer (BD Biosciences). All compensation and gating analyses were performed using FlowJo 10.5.3 (TreeStar, Ashland, OR, USA).

### Mitochondrial superoxide assay

Mitochondrial superoxide was detected by using a flow cytometry–based assay. Total PBMCs were washed with Hank’s balanced salt solution with calcium and magnesium (HBSS/Ca/Mg, Gibco, USA) to remove residual media by centrifugation. Cell pellet was stained with MitoSOX probe (Life Technologies, USA) at 37°C for 30 min following the manufacturer’s protocol. Cells were next washed with HBSS/Ca/Mg and incubated with antibodies to analyze CD14^+^ monocytes by flow cytometry. Cells fixed with 2% paraformaldehyde at room temperature for 30 min. Fluorescence intensity of the probe was then measured by flow cytometric analysis.

### Lipid peroxidation assay

Lipid peroxidation in total PBMC cultures was assessed by using the Click-iT Lipid Peroxidation Imaging Kit (Life Technologies, SA, USA) according to the manufacturer’s instructions. Cells were incubated with the LAA reagent (alkyne-modified linoleic acid) for detection of lipid peroxidation-derived protein modifications at 37°C for 1 h, and then washed with 1× DPBS. After cell centrifugation (1,500 rpm for 5 min) to remove the LAA reagent, cells were incubated with antibodies to measure lipid peroxidation in monocytes. Live/Dead staining was performed as described previously above and the cells fixed by adding cytofix/cytoperm (BD Bioscience, USA). After completing the LAA staining, fixed cells were washed, and resuspended in 1× DPBS, and then LAA fluorescence was analyzed by flow cytometry.

### Measurement of intracellular GSH levels

Intracellular GSH levels were measured in PBMC lysates by using a Glutathione Assay Kit (Cayman Chemical, USA) according to the manufacturer’s instructions. Total GSH levels were normalized by total protein concentration determined by using a Pierce BCA protein Assay kit (Thermo Fisher, IL, USA).

### Immunophenotyping by flow cytometry

PBMCs were stained with a 28-color flow cytometry panel modified from the optimized multicolor immunofluorescence panel (OMIP)-069 (104). Cells were incubated with LIVE/DEAD Fixable Blue Stain (Thermo Fischer, USA) and anti-CCR7 BV421 [Clone: G043H7] from BD Biosciences for 20 min at 37 °C, followed by an extracellular staining in PBS + 1%BSA with the following fluorochrome-conjugated antibodies: anti-CD57 Pacific Blue [Clone: HNK-1], anti-CD3 BV510 [Clone: SK7], anti-CD28 BV650 [Clone: CD28.2], anti-PD-1 BV785 [Clone: EH12.2H7], anti-CD14 Spark Blue 550 [Clone: 63D3], anti-CD45 PerCP [Clone: 2D1], anti-KLRG1 PE-Cy7 [Clone: 14C2A07], anti-CD19 Spark NIR 685 [Clone: HIB19], anti-CD38 APC/Fire810 [Clone: HIT2] from BioLegend; anti-CD45RA BUV395 [Clone: 5H9], anti-CD16 BUV496 [Clone: 3G8], anti-CD56 BUV737 [Clone: NCAM16.2], anti-CD8 BUV805 [Clone: SK1], anti-CD27 BV480 [Clone: O323], anti-CD127 APC-R700 [Clone: HIL-7R-M21], anti-HLA-DR APC-H7 [Clone: L243] from BD Biosciences; anti-CD20 cFluor V547 [Clone: 2H7], anti-CD4 cFluor yG584 [Clone: SK3] from Cytek; anti-KIR2D PE [Clone: NKVFS1], anti-KIR3DL1/DL2 PE [Clone: 5.133] from Miltenyi Biotech; anti-CD123 Super Bright 436 [Clone: 6H6], anti-TCRγδ PerCP-Vio700 [Clone: B1.1], anti-CD25 PE-Alexa Fluor 700 [Clone: CD25-3G10], CD87 APC [Clone: VIM5] from Thermo Fischer. Cells were fixed and permeabilized with Cytofix/Cytoperm (BD Biosciences, USA) then stained in cytoperm buffer (BD Biosciences, USA) with anti-Ki67 BV711 [Clone: B56] from BD Biosciences and anti-p16INK4a Alexa Fluor 647 [Clone: EPR1473] from Abcam. Data were acquired on Cytek Aurora flow cytometry (Cytek). Batch correction was performed on HLR-DR, CD127, CD45RA, CD16, CD56, CD8, CD27, CD3, CD28, CD25, CD45, TCRγγγγ, CD14, CD19, CD123, CD20, and CD4 using the R package, cyCombine, on the NIAID Skyline HPC cluster with R version 4.3.1. After transformation with an arcsine cofactor of 4000, the default parameters of cyCombine’s batch correct function were applied to each batch across experimental groups to preserve biological group variance while reducing batch variation during downstream analysis. Analysis was performed using FlowJo 10.5.3 (TreeStar, USA) and a cloud-based computational platform OMIQ.ai (Omiq, Santa Clara, CA). Equal sampling of 5,319 and 19,230 events per COVID and NC sample, respectively (lowest common denominator across samples) and a total of 500,000 events per group across all 120 samples were analyzed using uniform manifold approximation and projection (UMAP) clustering. We analyzed the abundance of the 25 clusters using EdgeR in OMIQ.ai between the COVID and NC samples.

### Single cell RNA sequencing analysis

PBMCs were fixed with the Evercode Cell Fixation v2 kit (Version 1.1, Parse Biosciences) following the manufacturer’s instructions. Approximately 500k to 1-million cells per sample were fixed and then frozen and stored at −80 °C until further processing. Fixed cells were then thawed and processed using the Evercode™ WT Mega v2 kit as per the manufacturer’s instructions (Parse Biosciences). In brief, cells were barcoded and 16-sublibraries were generated and sequenced on 2 lanes of NovaSeqX 25B, 150PE, flowcell to yield on average 52,000 reads per cell. FastQ files were aligned and analyzed using TrailmakerTM (Parse Biosciences, 2024). Overall, in 14-samples, 141,360 cells were sequenced with an average of 10,000 cells per sample. Single-cell RNA sequencing analyses were then performed using Seurat v5.1.0 (*95*) in R v4.3.3. Quality control was implemented by filtering out cells with more than 15% mitochondrial gene expression, indicative of low-quality cells, cells with fewer than 500 detected genes, suggestive of empty droplets, and cells with more than 10,000 detected genes, representative of doublets. The remaining cells were then normalized on an individual sample basis using the “SCTransform” function with default settings. A reciprocal PCA integration analysis was next performed to generate a single integrated dataset using the “IntegrateLayers” function with the “method” parameter set to “RPCAIntegration” and the “normalization.method” set to “SCT.” Dimensionality reduction was then conducted using the “RunPCA” function, followed by the “RunUMAP” function, utilizing the top 30 principal components. Cell type annotations were predicted using the “FindTransferAnchors” and “MapQuery” functions with the “PBMC CITE-seq” reference dataset (*95*). Finally, Gene Set Enrichment Analyses (GSEA) were performed using the fgsea v1.28.0 R package, based on average log2-fold changes obtained from the “FindMarkers” function with its “test.use” parameter set to “DESeq2” on pseudobulk profiles of each cell type.

### DNA methylation processing and analysis

Eighty-eight samples were processed on Infinium MethylationEPIC beadchips (v2.0, Illumina), containing just over 930,000 CpGs throughout the genome. ∼300ng of genomic DNA was bisulfite-converted using the EZ DNA Methylation-Lightning MagPrep kit (Zymo Research). Beta values were normalized using the SeSAMe openSesame function, implementing the Prep Code “CDPB”, which includes (C) infer channel for Infinium-I probes, (D) dye bias correction, (P) pOOBAH masking, and (B) noob background subtraction. Beta values and M-values were generated using the getBetas and BetaValueToMValue functions in SeSAMe. The EPICv2 design contains duplicate probes with differentiating suffixes appended to the CpG name. Duplicate probes were averaged and collapsed using the SeSAMe function betasCollapseToPfx. Quality control metrics were generated, and all samples had a successful detection fraction greater than 91%. Prior to differential methylation analysis, the following filtering criteria were implemented to remove probes: (1) CpGs with a detection P-value < 0.05 in more than 10% of samples, (2) CpGs whose probe included a SNP with a minor allele frequency > 0.05, (*96*) CpGs with cross-reactive probes. Differentially methylated CpGs were detected using the lmFit() function in the limma R package in conjunction with the treat() function. The model design compared COVID vs SARS-CoV-2-naïve controls and Recovered Individuals vs Long COVID, utilizing the makeContrasts() function. The topTreat function was used to adjust p-values using the FDR method to control the false discovery rate. CpGs were annotated using the Illumina EPICv2 annotation R package (IlluminaHumanMethylationEPICv2anno.20a1.hg38). DMRcate (v3.1.10) was used to evaluate evidence for differentially methylated regions of the genome. Gene ontology pathways were derived for significant CpGs using the gometh function in the missMethyl (v1.30.14) R package. Gene Ontology (GO) biological processes (BP), GO cellular components (CC), and GO molecular functions (MF) were evaluated for significant enrichment. Pathways from the Kyoto Encyclopedia of Genes and Genomes (KEGG) were also evaluated. The top 20 results with a p-value < 0.05 were extracted with the topGSA function. Using the genomic.features argument, we also evaluated enrichment of only significant CpGs located within promoter regions (TSS200, TSS1500, and 1stExon). The methylation age was predicted with the Horvath clock model using the agep() function in the wateRmelon R package, estimating the chronological age in years. The EPIC v2 design includes 345/353 CpGs from the Horvath 353 Model. After processing and filtering, there were 336-339 CpGs per sample used for age prediction.

### Statistical analyses

Statistical analyses were performed using non-parametric Mann-Whitney or Kruskal-Wallis test in GraphPad Prism 8.0.1 software (GraphPad, USA). Data are presented as median with interquartile ranges. Spearman’s correlation analyses were performed using R. Differences between groups or variables were considered significant when *p* < 0.05. Unsupervised clustering was performed on immunophenotyping data using FlowSom analysis of fcs files gated on CD45+ cells with the OMIQ software from Dotmatics (www.omiq.ai, www.dotmatics.com). Differential abundance analyses were used to generate volcano plots and heatmaps represent the relative expression of each immune marker. Elastinet regression models incorporating variables that demonstrated significant differences between groups was performed on both the biomarker and immunophenotyping datasets using glmnet in R (version 4.3.2). PCAs were generated using the packages FactoMineR and factoextra.

## Data Availability

All data produced in the present study are available upon reasonable request to the authors.

## Acknowledgments

The authors are grateful to the study participants enrolled in protocols for making this study possible. Authors also thank Dr Michael C. Sneller who gently provided the SARS-CoV-2-naïve control samples from the Longitudinal Study of COVID-19 Sequalae and Immunity (RECON-19) cohort. This research was supported [in part] by the Intramural Research Program of the National Institute of Allergy and Infectious Diseases (NIAID) as well as funded in part with federal funds from the NIAID, National Institutes of Health, Department of Health and Human Services under BCBB Support Services Contract HHSN316201300006W/75N93022F00001 to Guidehouse Digital.

## Funding

National Institutes of Health grant (HHSN261200800001E)

National Institutes of Health grant (HHSN2612015000031)

National Institutes of Health grant (75N910D00024)

MJP is supported on K23AI157875

PolyBioResearch Foundation

NIAID (R01AI141003/HHSN316201300006W/75N93022F00001)

NINDS (1R01NS136197)

## Author contributions

Conceptualization: SLL, KB, JMR and IS

Methodology: SLL, KB, JMR, AR, FXD, CM, LC, EPA, GR, JNM, TJH, MJP

Investigation: SLL, KB, JMR, TD, AE, RH

Visualization: SLL, KB, JMR, FXD, CM, EPA

Funding acquisition: IS, SGD, TJH, MJP

Supervision: IS, SGD, MJP

Writing – original draft: SLL, KB, JMR, IS

Writing – review & editing: SLL, KB, JMR, SGD, TJH, MJP, IS

## Supplementary Materials

### Supplementary figures

**Figure S1.** Phenotypic changes in circulating monocyte subsets stratified among groups.

**Figure S2.** Comparison of lipid mediators of inflammation in Long COVID.

**Figure S3.** Activation, senescence, and exhaustion status of PBMCs following COVID-19 infection.

**Figure S4.** Epigenetic vs chronological age in COVID-19.

### Supplementary tables

**Table S1.** Frequency of pre-existing conditions and hospitalization status in SARS-CoV-2 infected participants stratified by long COVID symptom status.

**Table S2.** Differential methylation analysis between COVID and NC.

**Table S3.** GO Pathway analysis.

## References and Notes

1. H. E. Davis, L. McCorkell, J. M. Vogel, E. J. Topol, Long COVID: major findings, mechanisms and recommendations. Nat Rev Microbiol 21, 133–146 (2023).

2. E. W. Ely, L. M. Brown, H. V. Fineberg, E. National Academies of Sciences, C. Medicine Committee on Examining the Working Definition for Long, Long Covid Defined. N Engl J Med 391, 1746–1753 (2024).

3. M. Cai, Y. Xie, E. J. Topol, Z. Al-Aly, Three-year outcomes of post-acute sequelae of COVID-19. Nat Med 30, 1564–1573 (2024).

4. L. Mateu et al., Determinants of the onset and prognosis of the post-COVID-19 condition: a 2-year prospective observational cohort study. Lancet Reg Health Eur 33, 100724 (2023).

5. M. J. Peluso, S. G. Deeks, Mechanisms of long COVID and the path toward therapeutics. Cell 187, 5500–5529 (2024).

6. H. K. Al-Hakeim, H. T. Al-Rubaye, A. F. Almulla, D. S. Al-Hadrawi, M. Maes, Chronic Fatigue, Depression and Anxiety Symptoms in Long COVID Are Strongly Predicted by Neuroimmune and Neuro-Oxidative Pathways Which Are Caused by the Inflammation during Acute Infection. J Clin Med 12, (2023).

7. H. K. Al-Hakeim, H. T. Al-Rubaye, D. S. Al-Hadrawi, A. F. Almulla, M. Maes, Long-COVID post-viral chronic fatigue and affective symptoms are associated with oxidative damage, lowered antioxidant defenses and inflammation: a proof of concept and mechanism study. Mol Psychiatry 28, 564–578 (2023).

8. S. L. Lage et al., Persistent Oxidative Stress and Inflammasome Activation in CD14(high)CD16(-) Monocytes From COVID-19 Patients. Front Immunol 12, 799558 (2021).

9. D. Sharma, T. D. Kanneganti, The cell biology of inflammasomes: Mechanisms of inflammasome activation and regulation. J Cell Biol 213, 617–629 (2016).

10. C. Vollbracht, K. Kraft, Oxidative Stress and Hyper-Inflammation as Major Drivers of Severe COVID-19 and Long COVID: Implications for the Benefit of High-Dose Intravenous Vitamin C. Front Pharmacol 13, 899198 (2022).

11. A. Sette, J. Sidney, S. Crotty, T Cell Responses to SARS-CoV-2. Annu Rev Immunol 41, 343–373 (2023).

12. P. Moss, The T cell immune response against SARS-CoV-2. Nat Immunol 23, 186–193 (2022).

13. K. Yin et al., Long COVID manifests with T cell dysregulation, inflammation and an uncoordinated adaptive immune response to SARS-CoV-2. Nat Immunol 25, 218–225 (2024).

14. K. Peppercorn, C. D. Edgar, T. Kleffmann, W. P. Tate, A pilot study on the immune cell proteome of long COVID patients shows changes to physiological pathways similar to those in myalgic encephalomyelitis/chronic fatigue syndrome. Sci Rep 13, 22068 (2023).

15. A. Mongelli et al., Evidence for Biological Age Acceleration and Telomere Shortening in COVID-19 Survivors. Int J Mol Sci 22, (2021).

16. X. Cao et al., Accelerated biological aging in COVID-19 patients. Nat Commun 13, 2135 (2022).

17. S. Lee et al., Virus-induced senescence is a driver and therapeutic target in COVID-19. Nature 599, 283–289 (2021).

18. M. Strioga, V. Pasukoniene, D. Characiejus, CD8+ CD28- and CD8+ CD57+ T cells and their role in health and disease. Immunology 134, 17–32 (2011).

19. J. Torres-Ruiz et al., Novel clinical and immunological features associated with persistent post-acute sequelae of COVID-19 after six months of follow-up: a pilot study. Infect Dis (Lond) 55, 243–254 (2023).

20. M. J. Peluso et al., Persistence, Magnitude, and Patterns of Postacute Symptoms and Quality of Life Following Onset of SARS-CoV-2 Infection: Cohort Description and Approaches for Measurement. Open Forum Infect Dis 9, ofab640 (2022).

21. M. Merad, J. C. Martin, Pathological inflammation in patients with COVID-19: a key role for monocytes and macrophages. Nat Rev Immunol 20, 355–362 (2020).

22. C. Junqueira et al., FcgammaR-mediated SARS-CoV-2 infection of monocytes activates inflammation. Nature 606, 576–584 (2022).

23. T. S. Rodrigues et al., Inflammasomes are activated in response to SARS-CoV-2 infection and are associated with COVID-19 severity in patients. J Exp Med 218, (2021).

24. A. J. Wilk et al., A single-cell atlas of the peripheral immune response in patients with severe COVID-19. Nat Med 26, 1070–1076 (2020).

25. J. Schulte-Schrepping et al., Severe COVID-19 Is Marked by a Dysregulated Myeloid Cell Compartment. Cell 182, 1419–1440 e1423 (2020).

26. B. K. Patterson et al., Persistence of SARS CoV-2 S1 Protein in CD16+ Monocytes in Post-Acute Sequelae of COVID-19 (PASC) up to 15 Months Post-Infection. Front Immunol 12, 746021 (2021).

27. C. Phetsouphanh et al., Immunological dysfunction persists for 8 months following initial mild-to-moderate SARS-CoV-2 infection. Nat Immunol 23, 210–216 (2022).

28. F. Bou-Abdallah, J. J. Paliakkara, G. Melman, A. Melman, Reductive Mobilization of Iron from Intact Ferritin: Mechanisms and Physiological Implication. Pharmaceuticals (Basel) 11, (2018).

29. E. Tyagi et al., Loss of p16(INK4A) stimulates aberrant mitochondrial biogenesis through a CDK4/Rb-independent pathway. Oncotarget 8, 55848–55862 (2017).

30. S. Saito et al., Metabolomic and immune alterations in long COVID patients with chronic fatigue syndrome. Front Immunol 15, 1341843 (2024).

31. S. M. R. Gomes et al., High levels of pro-inflammatory SARS-CoV-2-specific biomarkers revealed by in vitro whole blood cytokine release assay (CRA) in recovered and long-COVID-19 patients. PLoS One 18, e0283983 (2023).

32. H. Askari et al., A glance at the therapeutic potential of irisin against diseases involving inflammation, oxidative stress, and apoptosis: An introductory review. Pharmacol Res 129, 44–55 (2018).

33. S. Ryu et al., The matricellular protein SPARC induces inflammatory interferon-response in macrophages during aging. Immunity 55, 1609–1626 e1607 (2022).

34. L. J. H. Rasmussen, J. E. V. Petersen, J. Eugen-Olsen, Soluble Urokinase Plasminogen Activator Receptor (suPAR) as a Biomarker of Systemic Chronic Inflammation. Front Immunol 12, 780641 (2021).

35. T. Thomas, et al., COVID-19 infection alters kynurenine and fatty acid metabolism, correlating with IL-6 levels and renal status. JCI Insight 5, (2020).

36. Y. Cai et al., Kynurenic acid may underlie sex-specific immune responses to COVID-19. Sci Signal 14, (2021).

37. A. C. Wong et al., Serotonin reduction in post-acute sequelae of viral infection. Cell 186, 4851–4867 e4820 (2023).

38. V. P. Guntur et al., Signatures of Mitochondrial Dysfunction and Impaired Fatty Acid Metabolism in Plasma of Patients with Post-Acute Sequelae of COVID-19 (PASC). Metabolites 12, (2022).

39. V. Anantharaman, L. Aravind, Novel eukaryotic enzymes modifying cell-surface biopolymers. Biol Direct 5, 1 (2010).

40. L. Duca et al., The elastin receptor complex transduces signals through the catalytic activity of its Neu-1 subunit. J Biol Chem 282, 12484–12491 (2007).

41. J. B. Allard, C. Duan, IGF-Binding Proteins: Why Do They Exist and Why Are There So Many? Front Endocrinol (Lausanne) 9, 117 (2018).

42. Q. Zou et al., The CCCH-type zinc finger transcription factor Zc3h8 represses NF-kappaB-mediated inflammation in digestive organs in zebrafish. J Biol Chem 293, 11971–11983 (2018).

43. N. F. Meor Azlan, J. Zhang, Role of the Cation-Chloride-Cotransporters in Cardiovascular Disease. Cells 9, (2020).

44. E. Koulouridis, I. Koulouridis, Molecular pathophysiology of Bartter’s and Gitelman’s syndromes. World J Pediatr 11, 113–125 (2015).

45. D. Steel et al., Loss-of-Function Variants in HOPS Complex Genes VPS16 and VPS41 Cause Early Onset Dystonia Associated with Lysosomal Abnormalities. Ann Neurol 88, 867–877 (2020).

46. E. Monfrini et al., HOPS-associated neurological disorders (HOPSANDs): linking endolysosomal dysfunction to the pathogenesis of dystonia. Brain 144, 2610–2615 (2021).

47. X. Li et al., Structures and biological functions of zinc finger proteins and their roles in hepatocellular carcinoma. Biomark Res 10, 2 (2022).

48. D. Zhang et al., CIZ1 promoted the growth and migration of gallbladder cancer cells. Tumour Biol 36, 2583–2591 (2015).

49. G. Qiu, Y. Deng, ZFP64 transcriptionally activates PD-1 and CTLA-4 and plays an oncogenic role in esophageal cancer. Biochem Biophys Res Commun 622, 72–78 (2022).

50. B. Lu et al., A Transcription Factor Addiction in Leukemia Imposed by the MLL Promoter Sequence. Cancer Cell 34, 970–981 e978 (2018).

51. M. Zhu et al., Targeting ZFP64/GAL-1 axis promotes therapeutic effect of nab-paclitaxel and reverses immunosuppressive microenvironment in gastric cancer. J Exp Clin Cancer Res 41, 14 (2022).

52. Y. Cai et al., LncRNA CBR3-AS1 predicts a poor prognosis and promotes cervical cancer progression through the miR-3163/LASP1 pathway. Neoplasma 69, 1406–1417 (2022).

53. L. Xu et al., Upregulation of the long non-coding RNA CBR3-AS1 predicts tumor prognosis and contributes to breast cancer progression. Gene X 2, 100014 (2019).

54. M. Hou, N. Wu, L. Yao, LncRNA CBR3-AS1 potentiates Wnt/beta-catenin signaling to regulate lung adenocarcinoma cells proliferation, migration and invasion. Cancer Cell Int 21, 36 (2021).

55. Y. Guan, J. Yang, X. Liu, L. Chu, Long noncoding RNA CBR3 antisense RNA 1 promotes the aggressive phenotypes of non-small-cell lung cancer by sponging microRNA-509-3p and competitively upregulating HDAC9 expression. Oncol Rep 44, 1403–1414 (2020).

56. J. Cao, Q. Zhao, Q. Jia, Y. Li, LncRNA-CBR3-AS1 promotes and enhances the malignancy of ulcerative colitis via targeting miRNA-145-5p/FN1. Cell Mol Biol (Noisy-le-grand) 69, 181–186 (2023).

57. Y. Wang et al., LncRNA PlncRNA-1 participates in rheumatoid arthritis by regulating transforming growth factor beta1. Autoimmunity 53, 297–302 (2020).

58. D. M. Del Valle et al., An inflammatory cytokine signature predicts COVID-19 severity and survival. Nat Med 26, 1636–1643 (2020).

59. M. Jontvedt Jorgensen et al., Increased interleukin-6 and macrophage chemoattractant protein-1 are associated with respiratory failure in COVID-19. Sci Rep 10, 21697 (2020).

60. B. Wang et al., Metabolism pathways of arachidonic acids: mechanisms and potential therapeutic targets. Signal Transduct Target Ther 6, 94 (2021).

61. R. A. Lewis, K. F. Austen, R. J. Soberman, Leukotrienes and other products of the 5-lipoxygenase pathway. Biochemistry and relation to pathobiology in human diseases. N Engl J Med 323, 645–655 (1990).

62. M. Peters-Golden, W. R. Henderson, Jr., Leukotrienes. N Engl J Med 357, 1841–1854 (2007).

63. D. R. McDougle et al., Anti-inflammatory omega-3 endocannabinoid epoxides. Proc Natl Acad Sci U S A 114, E6034–E6043 (2017).

64. C. T. Bramante et al., Randomized Trial of Metformin, Ivermectin, and Fluvoxamine for Covid-19. N Engl J Med 387, 599–610 (2022).

65. D. Miklankova et al., Metformin Affects Cardiac Arachidonic Acid Metabolism and Cardiac Lipid Metabolite Storage in a Prediabetic Rat Model. Int J Mol Sci 22, (2021).

66. M. Maes, I. Mihaylova, J. C. Leunis, In chronic fatigue syndrome, the decreased levels of omega-3 poly-unsaturated fatty acids are related to lowered serum zinc and defects in T cell activation. Neuro Endocrinol Lett 26, 745–751 (2005).

67. S. J. Annesley, D. Missailidis, B. Heng, E. K. Josev, C. W. Armstrong, Unravelling shared mechanisms: insights from recent ME/CFS research to illuminate long COVID pathologies. Trends Mol Med 30, 443–458 (2024).

68. M. J. Peluso, M. R. Hanson, S. G. Deeks, Infection-associated chronic conditions: Why Long Covid is our best chance to untangle Osler’s web. Sci Transl Med 16, eado2101 (2024).

69. B. Kavyani et al., Could the kynurenine pathway be the key missing piece of Myalgic Encephalomyelitis/Chronic Fatigue Syndrome (ME/CFS) complex puzzle? Cell Mol Life Sci 79, 412 (2022).

70. M. A. Kirby, S. Weitzman, M. H. Freedman, Juvenile chronic myelogenous leukemia: differentiation from infantile cytomegalovirus infection. Am J Pediatr Hematol Oncol 12, 292–296 (1990).

71. H. G. Herrod, L. W. Dow, J. L. Sullivan, Persistent epstein-barr virus infection mimicking juvenile chronic myelogenous leukemia: immunologic and hematologic studies. Blood 61, 1098–1104 (1983).

72. S. Yetgin, M. Cetin, I. Yenicesu, F. Ozaltin, D. Uckan, Acute parvovirus B19 infection mimicking juvenile myelomonocytic leukemia. Eur J Haematol 65, 276–278 (2000).

73. E. Vadillo et al., A Shift Towards an Immature Myeloid Profile in Peripheral Blood of Critically Ill COVID-19 Patients. Arch Med Res 52, 311–323 (2021).

74. E. M. Pietras et al., Chronic interleukin-1 exposure drives haematopoietic stem cells towards precocious myeloid differentiation at the expense of self-renewal. Nat Cell Biol 18, 607–618 (2016).

75. M. J. Peluso et al., Plasma-based antigen persistence in the post-acute phase of COVID-19. Lancet Infect Dis, (2024).

76. Z. Swank et al., Persistent Circulating Severe Acute Respiratory Syndrome Coronavirus 2 Spike Is Associated With Post-acute Coronavirus Disease 2019 Sequelae. Clin Infect Dis 76, e487–e490 (2023).

77. Z. Swank et al., Measurement of circulating viral antigens post-SARS-CoV-2 infection in a multicohort study. Clin Microbiol Infect 30, 1599–1605 (2024).

78. M. J. Peluso et al., Chronic viral coinfections differentially affect the likelihood of developing long COVID. J Clin Invest 133, (2023).

79. J. Klein et al., Distinguishing features of long COVID identified through immune profiling. Nature 623, 139–148 (2023).

80. K. Pinchaud et al., Impact of Dietary Arachidonic Acid on Gut Microbiota Composition and Gut-Brain Axis in Male BALB/C Mice. Nutrients 14, (2022).

81. D. Zheng, T. Liwinski, E. Elinav, Inflammasome activation and regulation: toward a better understanding of complex mechanisms. Cell Discov 6, 36 (2020).

82. R. Izzo et al., Combining L-Arginine with vitamin C improves long-COVID symptoms: The LINCOLN Survey. Pharmacol Res 183, 106360 (2022).

83. V. Bronte et al., Baricitinib restrains the immune dysregulation in patients with severe COVID-19. J Clin Invest 130, 6409–6416 (2020).

84. S. A. Lee et al., Impact of HIV on CD8+ T cell CD57 expression is distinct from that of CMV and aging. PLoS One 9, e89444 (2014).

85. H. Rayess, M. B. Wang, E. S. Srivatsan, Cellular senescence and tumor suppressor gene p16. Int J Cancer 130, 1715–1725 (2012).

86. D. K. J. Pieren et al., Regulatory KIR(+) RA(+) T cells accumulate with age and are highly activated during viral respiratory disease. Aging Cell 20, e13372 (2021).

87. A. Chalkias et al., Circulating suPAR associates with severity and in-hospital progression of COVID-19. Eur J Clin Invest 52, e13794 (2022).

88. J. B. Mannick, D. W. Lamming, Targeting the biology of aging with mTOR inhibitors. Nat Aging 3, 642–660 (2023).

89. X. Xu, L. Ye, K. Araki, R. Ahmed, mTOR, linking metabolism and immunity. Semin Immunol 24, 429–435 (2012).

90. Y. Ma-Lauer et al., p53 down-regulates SARS coronavirus replication and is targeted by the SARS-unique domain and PLpro via E3 ubiquitin ligase RCHY1. Proc Natl Acad Sci U S A 113, E5192–5201 (2016).

91. K. Bhardwaj, P. Liu, J. L. Leibowitz, C. C. Kao, The coronavirus endoribonuclease Nsp15 interacts with retinoblastoma tumor suppressor protein. J Virol 86, 4294–4304 (2012).

92. N. Singh, A. Bharara Singh, S2 subunit of SARS-nCoV-2 interacts with tumor suppressor protein p53 and BRCA: an in silico study. Transl Oncol 13, 100814 (2020).

93. G. Saini, R. Aneja, Cancer as a prospective sequela of long COVID-19. Bioessays 43, e2000331 (2021).

94. A. M. Evans, C. D. DeHaven, T. Barrett, M. Mitchell, E. Milgram, Integrated, nontargeted ultrahigh performance liquid chromatography/electrospray ionization tandem mass spectrometry platform for the identification and relative quantification of the small-molecule complement of biological systems. Anal Chem 81, 6656–6667 (2009).

95. Y. Hao et al., Dictionary learning for integrative, multimodal and scalable single-cell analysis. Nat Biotechnol 42, 293–304 (2024).

96. Y. Hao et al., Integrated analysis of multimodal single-cell data. Cell 184, 3573–3587 e3529 (2021).

