## Supplemental Material for "Persistent immune dysregulation and metabolic alterations following SARS-CoV-2 infection"

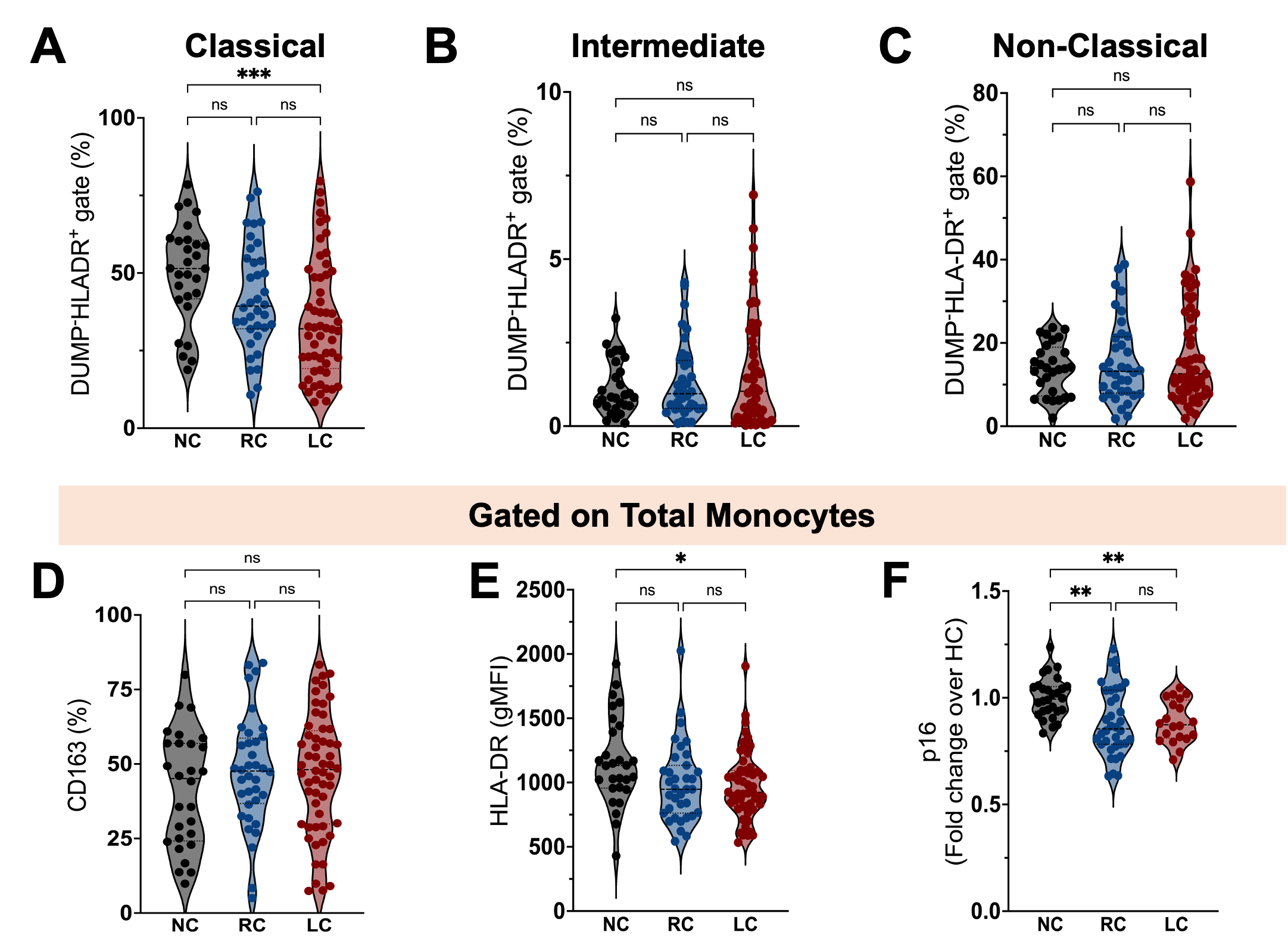

**Supplemental Figure 1. Phenotypic changes in circulating monocyte subsets stratified among groups.** Percentage of classical **(A)**, intermediate **(B)** and non-classical **(C)** monocytes among circulating mononuclear myeloid cells (HLADR+CD2−CD3−CD19−CD20−CD56−CD66−) in SARS-CoV-2-naive controls (NC, n=28) and COVID-19 participants stratified into COVID-19 recovered (RC, n=36) or presenting with long COVID symptoms (LC, n=56). Percentages of CD163^+^ cells **(D)**, and geometric mean fluorescence intensity (gMFI) expression of HLA-DR **(E)** and p16^INK4a^ **(F)** on circulating monocytes from NC, RC and LC. gMFI data are expressed as median of the fold change (gMFI of each sample over the gMFI mean of the NC group). Bars represent median ± quartiles. Statistical analysis was performed using a two-sided Mann-Whitney test (*, *P* < 0.05, **, *P* < 0.005, ***, *P* < 0.0005).

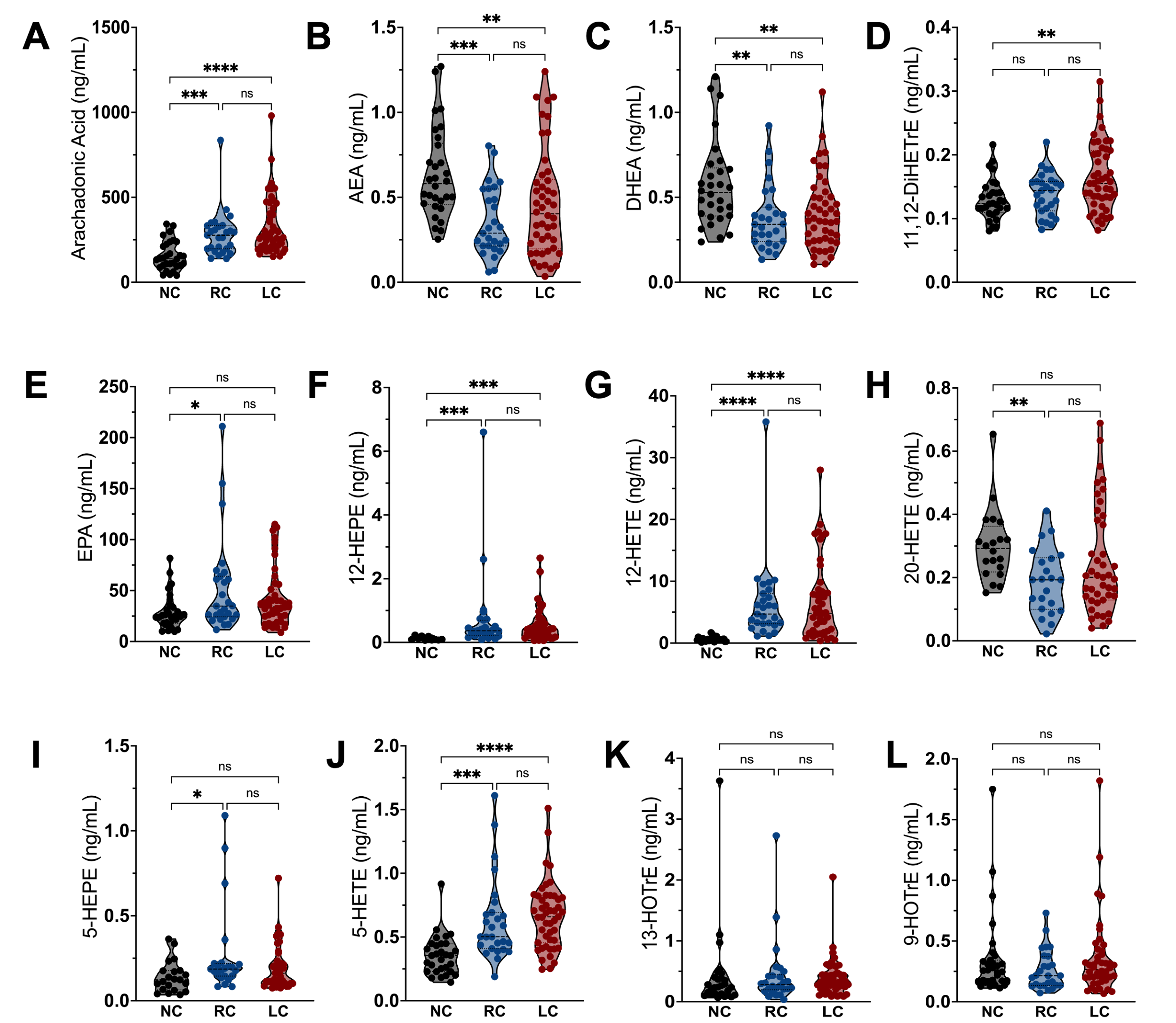

**Supplemental Figure 2. Comparison of lipid mediators of inflammation in Long COVID.** Cellular levels of (**A)** arachidonic acid (AA), (**B**) (AEA), (**C**) docosahexaenoyl ethanolamide (DHEA), (**D**) 11,12- dihydroxyeicosatreinoic acid (DiHETrE), (**E**) eicosapentaenoic acid (EPA), (**F**) 12- hydroxyeicosapentaenoic acid (HEPE), (**G**) hydroxyeicosatetraenoic acid (HETE), (**H**) 20-HETE, (**I**) 5-HEPE, (**J**) 5-HETE, (**K**) 13- hydroxyoctadecatrienoic acid (HOTrE), and (**L**) 9-HOTrE in SARS-CoV-2 unexposed controls (NC, n=28) and COVID-19 participants stratified into COVID-19 recovered individuals (RC, n=36) and participants presenting with long COVID symptoms (LC, n=56). Bars represent median ± quartiles. Statistical analysis was performed using a Kruskal-Wallis test (*, *P* < 0.05, **, *P* < 0.005, ***, *P* < 0.0005, **** *P* < 0.0001).

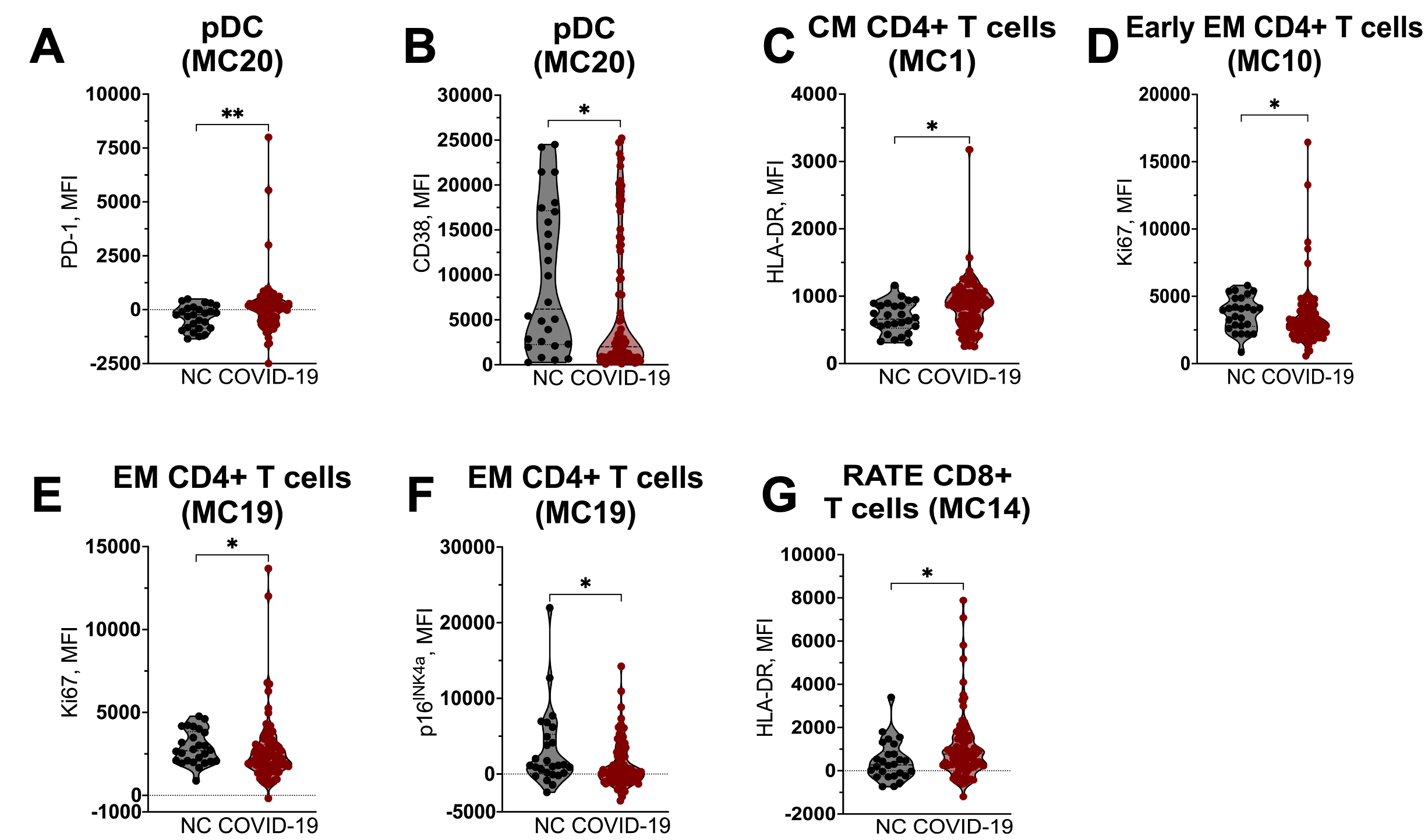

**Supplemental Figure 3. Activation, senescence, and exhaustion status of PBMCs following COVID-19 infection.** Mean fluorescent intensity (MFI) expression of (**A**) PD-1 and (**B**) CD38 in pDCs (MC20), (**C**) HLA-DR in central memory (42) CD4 T-cells (MC01), (**D**) Ki67 in early effector memory (EM) CD4 T cells, (**E**) Ki67 and p16^INK4a^ in EM CD4 T cells, and (**G**) HLA-DR in CD45RA+ terminal effector (RATE) CD8 T-cells in SARS-CoV-2 unexposed controls (NC compared to COVID-19 patients. Bars represent median ± quartiles. Statistical analysis was performed using a two-sided Mann-Whitney test (*, *P* < 0.05, **, *P* < 0.005).

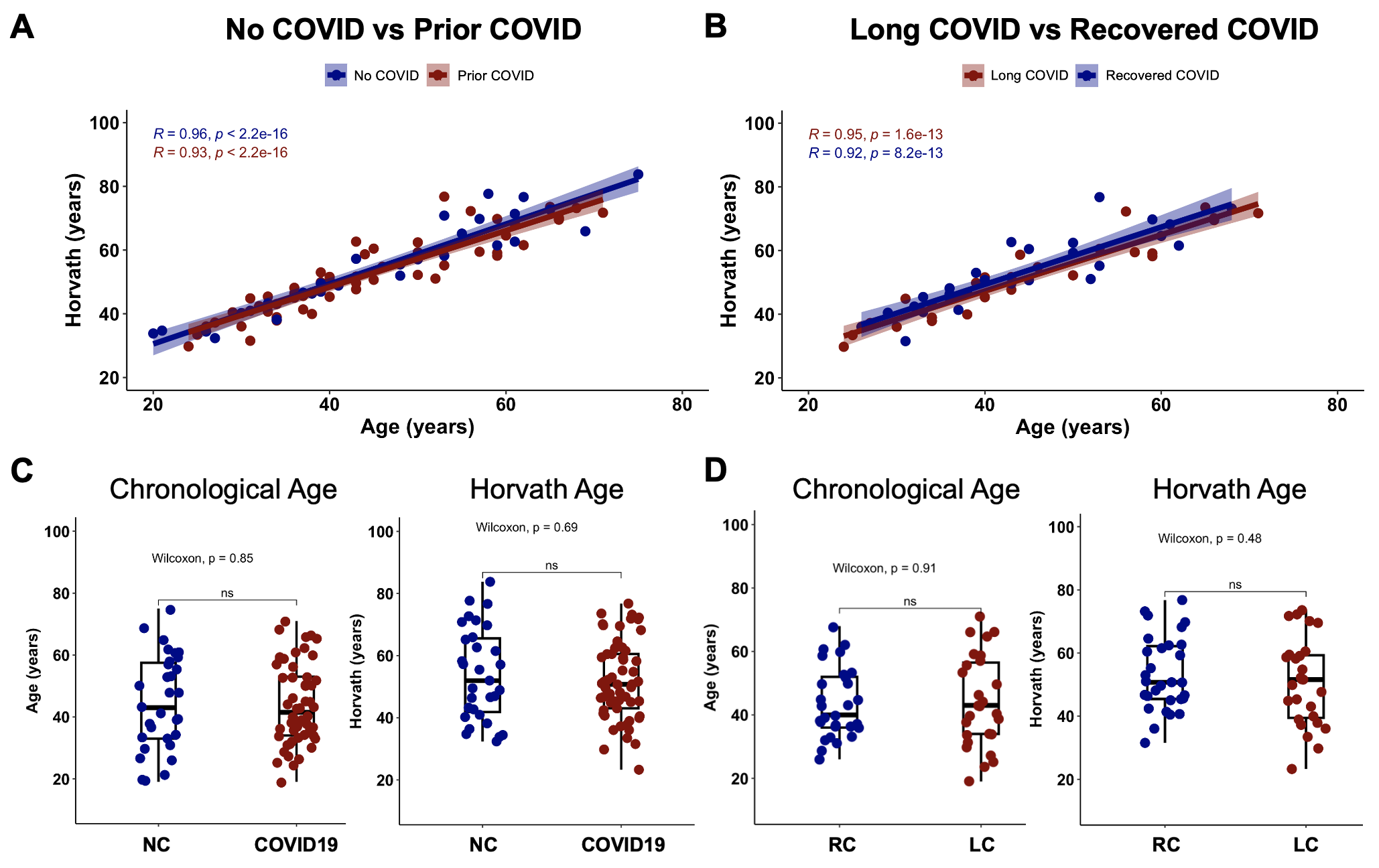

**Supplemental Figure 4. Epigenetic vs chronological age in COVID-19.** Correlations of chronological age and Horvath epigenetic clock in (**A**) SARS-CoV-2-naive controls (NC) compared to participants with prior COVID-19 and (**B**) between recovered COVID-19 (RC) and Long COVID (LC) participants. Comparisons of chronological and epigenetic age in (**C**) NC and COVID-19 and (**D**) RC and LC patients. Bars represent median ± quartiles. Statistical analysis was performed using a Wilcoxon test.

| **Characteristic** | | **COVID-19 (*n = 96*)** | **RC (*n = 38*)** | **LC (*n = 58*)** |
| --- | --- | --- | --- | --- |
| **Pre-Existing Condition** | | 35 (36.5%) | 11 (28.9%) | 16 (27.6%) |
|  | *Autoimmune Disease* | *7 (7.3%)* | *1 (2.6%)* | *6 (15.8%)* |
|  | *Cancer* | *3 (3.1%)* | *1 (2.6%)* | *2 (5.3%)* |
|  | *Diabetes* | *11 (11.5%)* | *5 (13.2%)* | *6 (15.8%)* |
|  | *Hypertension* | *13 (13.5%)* | *3 (7.9%)* | *10 (26.3%)* |
|  | *Lung Disease* | *17 (17.7%)* | *6 (15.8%)* | *11 (28.9%)* |
| **Days from first symptom** | | 122.5 (range: 91-245) | 119.4 (range: 91 - 142) | 124.6 (range: 92 - 245) |
| **Hospitalization** | | 23 (24.0%) | 7 (18.4%) | 16 (27.6%) |
|  | *Oxygen* | *19 (82.6%)* | *5 (71.4%)* | *14 (87.5%)* |
|  | *ICU* | *7 (30.4%)* | *1 (14.3%)* | *6 (37.5%)* |
|  | *Ventilator* | *2 (8.7%)* | *1 (14.3%)* | *1 (6.3%)* |

**Supplemental Table 1. Frequency of pre-existing conditions and hospitalization status in SARS-CoV-2 infected participants stratified by long COVID symptom status.** RC, recovered COVID-19; LC, long COVID symptoms.

| **Experimental Group** | **CpG** | **Gene** | **Function** | **Reference** |
| --- | --- | --- | --- | --- |
| Highly methylated in COVID-19 | cg11450715 | *ERICH2* | Poorly characterized transcription factor |  |
|  | cg22438603 | *SLC12A* | Encode cation-chloride membrane transport proteins | (*1, 2*) |
|  |  |  | Regulate physiological processes including cell volume, blood pressure, neuronal action potential |  |
|  |  |  | Mutations can cause Gitelman's syndrome (muscle weakness, muscle cramps, fatigue, dizziness) | (*3*) |
|  |  |  | Functional role in pathophysiology of hypertension and cardiovascular disease | (*4*) |
|  | cg10729317 | *PCED1A* | Member of the GDSL/SGNH superfamily of hydrolytic enzymes, esterase and lipase activity | (*5*) |
|  |  | *VP16* | Encodes protein involved in autophagosome-lysosome fusion | (*6, 7*) |
|  |  |  | Mutations have been linked to dystonia, neurological movement disorder |  |
|  | cg03927520 | *GLB1L* | Predicted to enable beta-galactosidase activity, which acts in carbohydrate catabolic process. Variants leading to β‐galactosidase deficiency can cause metabolic disorders: Morquio B and GM1‐gangliosidosis (severe muscle weakness and atrophy) | (*8*) |
|  |  | *STK16* | Ubiquitosly expressed serine/threonine protein kinase regulates actin dynamics to maintain Golgi structure and control cell cycle progression | (*9*) |
|  | cg27242549 | *ZC3H8* | Member of the zinc finger family of transcription factors, protects hepatocytes from degeneration in zebrafish by inhibiting inflammation and NFκB signaling | (*10*) |
| Highly methylated in NC | cg22805491 | *ZFP64* | Positively regulates development of multiple cancers including hepatocellular carcinoma and esophageal cancer | (*11-13*) |
|  |  |  | Activates the promoter of mixed spectrum leukemia oncogene as a positive feedback loop | (*14*) |
|  |  |  | Promotor of Gal-1, PD-1, and CTLA-4 genes | (*13, 15*) |
|  | cg17499729 | *CBR3-AS1* | Oncogenic long non-coding RNA in several cancers | (*16-20*) |
|  |  |  | Participates in chronic inflammatory and autoimmune diseases including ulcerative colitis and rheumatoid arthritis | (*21*) |
|  |  |  | Reportedly downregulated in colorectal cancer | (*22*) |
|  |  | *RPS9* | Component of the 40S subunit of ribosomes found highly expressed in myeloma cells | (*23*) |
|  | cg10438232 | *CDON* | Encodes a member of a cell-surface receptor complex that mediates cell-cell interactions and positively regulates myogenesis | (*24*) |
|  |  |  | Involved in prostate tumor cell growth and invasion and lung cancer cell proliferation and tumorigenicity | (*25, 26*) |
|  |  |  | Tumor suppressor in neuroblastomas | (*27*) |
|  | cg10165071 | *IGFBP* | Regulation of somatic growth and cellular proliferation by interacting with insulin-like growth factors (IGFs) | (*28*) |
|  | cg25075434 | *KIZ* | Centrosome stabilization and protein kinase binding. Overexpression leads to down-modulation of IL-1β-induced NF-κB signaling by targeting TRAF6 | (*29*) |

**Supplemental Table 2. Differential methylation analysis between COVID and NC.** Associated genes and function of differentially methylated CpGs between COVID-19 and NC individuals.

| **Gene** | **Ontology** | **Term** | **Genes (n)** | **DE (n)** | **P-Value** | **FDR** |
| --- | --- | --- | --- | --- | --- | --- |
| GO:0005654 | CC | nucleoplasm | 3966 | 2426 | 2.67E-35 | 5.96E-31 |
| GO:0005634 | CC | nucleus | 7442 | 4135 | 6.64E-32 | 7.40E-28 |
| GO:0031981 | CC | nuclear lumen | 4639 | 2652 | 1.61E-27 | 1.20E-23 |
| GO:0043231 | CC | intracellular membrane-bounded organelle | 11779 | 6324 | 4.91E-25 | 2.73E-21 |
| GO:0043229 | CC | intracellular organelle | 12843 | 6867 | 2.94E-24 | 1.31E-20 |
| GO:0031974 | CC | membrane-enclosed lumen | 5728 | 3188 | 1.25E-23 | 3.48E-20 |
| GO:0043233 | CC | organelle lumen | 5728 | 3188 | 1.25E-23 | 3.48E-20 |
| GO:0070013 | CC | intracellular organelle lumen | 5728 | 3188 | 1.25E-23 | 3.48E-20 |
| GO:0140513 | CC | nuclear protein-containing complex | 1195 | 785 | 1.94E-23 | 4.80E-20 |
| GO:1902494 | CC | catalytic complex | 1649 | 1038 | 1.23E-22 | 2.74E-19 |
| GO:0005622 | CC | intracellular anatomical structure | 14293 | 7570 | 1.32E-19 | 2.66E-16 |
| GO:0044238 | BP | primary metabolic process | 9769 | 5199 | 2.83E-19 | 5.25E-16 |
| GO:0097159 | MF | organic cyclic compound binding | 5898 | 3260 | 1.43E-18 | 2.45E-15 |
| GO:0071840 | BP | cellular component organization or biogenesis | 6548 | 3733 | 2.94E-18 | 4.68E-15 |
| GO:0032991 | CC | protein-containing complex | 5600 | 3089 | 4.11E-18 | 6.11E-15 |
| GO:1990234 | CC | transferase complex | 833 | 549 | 7.83E-18 | 1.09E-14 |
| GO:0034654 | BP | nucleobase-containing compound biosynthetic process | 4710 | 2580 | 9.83E-18 | 1.29E-14 |
| GO:0043227 | CC | membrane-bounded organelle | 12861 | 6782 | 1.14E-17 | 1.42E-14 |
| GO:0006139 | BP | nucleobase-containing compound metabolic process | 5638 | 3047 | 1.89E-17 | 2.21E-14 |
| GO:0006325 | BP | chromatin organization | 987 | 644 | 9.37E-17 | 1.04E-13 |

**Supplemental Table 3. GO Pathway analysis.** CC, cellular component; BP, biological process; MF, molecular function. DE, number of genes that are differentially methylated in pathway.

**Supplemental References and Notes**

1.	S. Zhang *et al.*, The role of SLC12A family of cation-chloride cotransporters and drug discovery methodologies. *J Pharm Anal* **13**, 1471-1495 (2023).

2.	J. P. Arroyo, K. T. Kahle, G. Gamba, The SLC12 family of electroneutral cation-coupled chloride cotransporters. *Mol Aspects Med* **34**, 288-298 (2013).

3.	E. Koulouridis, I. Koulouridis, Molecular pathophysiology of Bartter's and Gitelman's syndromes. *World J Pediatr* **11**, 113-125 (2015).

4.	N. F. Meor Azlan, J. Zhang, Role of the Cation-Chloride-Cotransporters in Cardiovascular Disease. *Cells* **9**, (2020).

5.	V. Anantharaman, L. Aravind, Novel eukaryotic enzymes modifying cell-surface biopolymers. *Biol Direct* **5**, 1 (2010).

6.	D. Steel *et al.*, Loss-of-Function Variants in HOPS Complex Genes VPS16 and VPS41 Cause Early Onset Dystonia Associated with Lysosomal Abnormalities. *Ann Neurol* **88**, 867-877 (2020).

7.	E. Monfrini *et al.*, HOPS-associated neurological disorders (HOPSANDs): linking endolysosomal dysfunction to the pathogenesis of dystonia. *Brain* **144**, 2610-2615 (2021).

8.	J. J. Pedersen *et al.*, beta-Galactosidase deficiency in the GLB1 spectrum of lysosomal storage disease can present with severe muscle weakness and atrophy. *JIMD Rep* **63**, 540-545 (2022).

9.	J. Liu *et al.*, STK16 regulates actin dynamics to control Golgi organization and cell cycle. *Sci Rep* **7**, 44607 (2017).

10.	Q. Zou *et al.*, The CCCH-type zinc finger transcription factor Zc3h8 represses NF-kappaB-mediated inflammation in digestive organs in zebrafish. *J Biol Chem* **293**, 11971-11983 (2018).

11.	X. Li *et al.*, Structures and biological functions of zinc finger proteins and their roles in hepatocellular carcinoma. *Biomark Res* **10**, 2 (2022).

12.	D. Zhang *et al.*, CIZ1 promoted the growth and migration of gallbladder cancer cells. *Tumour Biol* **36**, 2583-2591 (2015).

13.	G. Qiu, Y. Deng, ZFP64 transcriptionally activates PD-1 and CTLA-4 and plays an oncogenic role in esophageal cancer. *Biochem Biophys Res Commun* **622**, 72-78 (2022).

14.	B. Lu *et al.*, A Transcription Factor Addiction in Leukemia Imposed by the MLL Promoter Sequence. *Cancer Cell* **34**, 970-981 e978 (2018).

15.	M. Zhu *et al.*, Targeting ZFP64/GAL-1 axis promotes therapeutic effect of nab-paclitaxel and reverses immunosuppressive microenvironment in gastric cancer. *J Exp Clin Cancer Res* **41**, 14 (2022).

16.	Y. Cai *et al.*, LncRNA CBR3-AS1 predicts a poor prognosis and promotes cervical cancer progression through the miR-3163/LASP1 pathway. *Neoplasma* **69**, 1406-1417 (2022).

17.	L. Xu *et al.*, Upregulation of the long non-coding RNA CBR3-AS1 predicts tumor prognosis and contributes to breast cancer progression. *Gene X* **2**, 100014 (2019).

18.	Y. Guan, J. Yang, X. Liu, L. Chu, Long noncoding RNA CBR3 antisense RNA 1 promotes the aggressive phenotypes of non‑small‑cell lung cancer by sponging microRNA‑509‑3p and competitively upregulating HDAC9 expression. *Oncol Rep* **44**, 1403-1414 (2020).

19.	M. Hou, N. Wu, L. Yao, LncRNA CBR3-AS1 potentiates Wnt/beta-catenin signaling to regulate lung adenocarcinoma cells proliferation, migration and invasion. *Cancer Cell Int* **21**, 36 (2021).

20.	J. Cao, Q. Zhao, Q. Jia, Y. Li, LncRNA-CBR3-AS1 promotes and enhances the malignancy of ulcerative colitis via targeting miRNA-145-5p/FN1. *Cell Mol Biol (Noisy-le-grand)* **69**, 181-186 (2023).

21.	Y. Wang *et al.*, LncRNA PlncRNA-1 participates in rheumatoid arthritis by regulating transforming growth factor beta1. *Autoimmunity* **53**, 297-302 (2020).

22.	M. Yang *et al.*, Long Non-coding RNA CBR3 Antisense RNA 1 is Downregulated in Colorectal Cancer and Inhibits miR-29a-Mediated Cell Migration and Invasion. *Mol Biotechnol* **64**, 773-779 (2022).

23.	J. Kang *et al.*, Ribosomal proteins and human diseases: molecular mechanisms and targeted therapy. *Signal Transduct Target Ther* **6**, 323 (2021).

24.	L. Sanchez-Arrones, M. Cardozo, F. Nieto-Lopez, P. Bovolenta, Cdon and Boc: Two transmembrane proteins implicated in cell-cell communication. *Int J Biochem Cell Biol* **44**, 698-702 (2012).

25.	T. Hayashi *et al.*, Identification of transmembrane protein in prostate cancer by the Escherichia coli ampicillin secretion trap: expression of CDON is involved in tumor cell growth and invasion. *Pathobiology* **78**, 277-284 (2011).

26.	Y. E. Leem, H. L. Ha, J. H. Bae, K. H. Baek, J. S. Kang, CDO, an Hh-coreceptor, mediates lung cancer cell proliferation and tumorigenicity through Hedgehog signaling. *PLoS One* **9**, e111701 (2014).

27.	B. Gibert *et al.*, Regulation by miR181 family of the dependence receptor CDON tumor suppressive activity in neuroblastoma. *J Natl Cancer Inst* **106**, (2014).

28.	P. F. Collett-Solberg, P. Cohen, The role of the insulin-like growth factor binding proteins and the IGFBP proteases in modulating IGF action. *Endocrinol Metab Clin North Am* **25**, 591-614 (1996).

29.	J. Sun, Q. Yang, E. Liu, D. Chen, Q. Sun, KIZ/GM114 Balances the NF-kB Signaling by Antagonizing the Association of TRAF2/6 With Their Upstream Adaptors. *Front Cell Dev Biol* **10**, 877039 (2022).
